# Shared genetic architecture between mental health and the brain functional connectome in the UK Biobank

**DOI:** 10.1101/2022.06.24.22276846

**Authors:** Daniel Roelfs, Oleksandr Frei, Dennis van der Meer, Elleke Tissink, Alexey Shadrin, Dag Alnæs, Ole A. Andreassen, Lars T. Westlye, Tobias Kaufmann

## Abstract

Psychiatric disorders are complex clinical conditions with large heterogeneity and overlap in symptoms, genetic liability and brain imaging abnormalities. Building on a dimensional conceptualization of mental health, previous studies have reported genetic overlap between psychiatric disorders and population-level mental health, and between psychiatric disorders and brain functional connectivity. Here, in 30.701 participants aged 45-82 from the UK Biobank we map the genetic associations between self-reported mental health and resting-state fMRI-based measures of brain network function. Multivariate Omnibus Statistical Test revealed 10 genetic loci associated with population-level mental symptoms. Next, conjunctional FDR identified 23 shared genetic variants between these symptom profiles and fMRI-based brain network measures. Functional annotation implicated genes involved in brain structure and function, in particular synaptic processes. These findings provide further genetic evidence of an association between brain function and mental health traits in the population.

## Introduction

Psychiatric disorders are complex, with a polygenic architecture, and large degree of overlapping symptoms and risk factors. Both imaging and genetics studies have shown numerous but small associations between brain phenotypes, psychiatric disorders, and genetics, such as schizophrenia (SCZ)^1– 3^, bipolar disorder (BIP)^4,5^, major depressive disorder (MDD)^6,7^, and anxiety disorder (ANX)^8–10^. Interactions between various brain phenotypes and genetics have been reported across structural^11,12^ and functional^2,7,10^ imaging modalities.

We have recently deployed a multivariate analysis to study the genetic architecture of brain functional connectivity, revealing genetic variants associated with functional brain connectivity as well as variance in brain activity over time^13^. The results showed meaningful overlap with psychiatric disorders, pointing at synapse-related pathways among the biological processes shared between disorders and brain function^13^.

Previous studies have shown widespread phenotypic and genetic overlap between psychiatric disorders^14– 18^. In addition, patients within a diagnostic category can display a wide variety of symptoms. This heterogeneity complicates both diagnosis and therapeutic response due to overlapping symptoms and generally low specificity of diagnostic features^19,20^. While the mental health of any individual in the population varies over the course of a lifetime, most will not meet diagnostic criteria for a psychiatric disorder^21,22^. In order to capture the variance encompassing psychiatric symptoms that is lacking in traditional case-control studies, one can use population-level mental health questionnaires as implemented in the UK Biobank^23^. This facilitates analyses using the continuous scales which enable data-driven clustering methods to extract different profiles each capturing a separate domain relevant to mental health in a sample without individuals diagnosed with a psychiatric disorder, taking advantage of larger sample sizes. Using independent component analysis (ICA), we have previously derived 13 mental health profiles from UK Biobank data, and showed that, although phenotypically independent (by design) they nonetheless share genetic underpinnings24.

Here, we aimed to uncover the genetic architecture of mental symptoms and identify shared genetic loci with neurobiological processes related to brain function. Using multivariate analysis^25^, we generated multivariate genome-wide association statistics across our previously identified 13 population-level mental health profiles^24^. This allowed us to identify new gene variants associated with mental health symptoms and traits such as psychosis, depression, and anxiety in the UK Biobank sample not captured in a univariate analysis. Further, we combined this multivariate genetic profile of mental health with GWAS summary statistics of 7 psychiatric disorders and with our previously identified multivariate profiles of functional brain connectivity and variance in brain activity over time^24^. This research aims to provide insight into the biological underpinnings of mental health symptoms.

## Methods

### Sample and exclusion criteria

We utilized data from the UK Biobank^26^ with permission no. 27412. All participants provided signed informed consent before inclusion in the study. The UK Biobank was approved by the National Health Service National Research Ethics Service (ref. 11/NW/0382). We previously used data from the online follow-up questionnaire on mental health to define 13 phenotypically independent profiles relevant for mental health^24^. In addition, we also utilized imaging data provided by the UK Biobank, the procedure of processing and analyzing the imaging data is described a previous paper^13^. For this study we deployed the summary statistics from these previous studies.

### Image acquisition and pre-processing

The processing pipeline for imaging data used for the multivariate GWAS is described in Roelfs *et al*.^13^. In short, images were acquired using 3T Siemens Magnetom Skyra scanners with a 32 channel head coil (Siemens Healthcare GmbH, Erlangen, Germany) at four different sites in the UK. The fMRI data was recorded using a gradient-echo echo planar imaging sequence with x8 multislice acceleration (TR: 0.735s, TE: 39ms, FOV: 88×88×64 matrix, FA: 52°) with a voxel size of 2.4×2.4×2.4mm. Data is processed by the UK Biobank team following the protocol described in Alfaro-Almagro *et al*.^27^.

### Multivariate Genome-Wide Analysis

In this study we applied the Multivariate Omnibus Statistical Test (MOSTest) to the phenotypic data from the ICA decomposition described in Roelfs *et al*.^24^. MOSTest deploys the univariate test-statistics for each SNP and computes a multivariate test statistic through single random permutations of the genotype vector. This method is described in Van der Meer *et al* ^25^. We performed positional gene mapping using Functional Mapping and Annotation (FUMA)^28^. We also used a built-in tool to follow up these gene mapping analyses using MAGMA to connect the identified genes with tissue types^29^. We analyzed gene sets using the reactome toolbox^30^ to identify biological processes associated with the genes associated with the summary statistics identified by FUMA.

### Pleiotropy-informed conjunctional false discovery rate

In order to quantify the degree of genetic overlap and identify shared genetic loci between the mental health profiles and the imaging features we deployed the pleiotropy-informed conjunctional false discovery rate (conjFDR) through the pleioFDR toolbox^31^. One of the advantages of conjFDR is that it can identify shared genetic loci regardless of effect direction and effect size, a feature that is useful when working with multivariate measures where effect direction might be lacking.

## Results

MOSTest revealed 10 significant loci across the 13 previously identified profiles of mental health (Figure 1). FUMA and its positional mapping tool revealed 48 genes associated with these loci (See Suppl. Table 1), that were linked by MAGMA to a number of brain structures such as the cerebellum and amygdala (Suppl. Figure 1). Among the identified genes, those mapped from the strongest GWAS loci were *ADH1B* and *ADH5* (chromosome 4) and *CRHR1* (chromosome 17).

**Figure 1.**
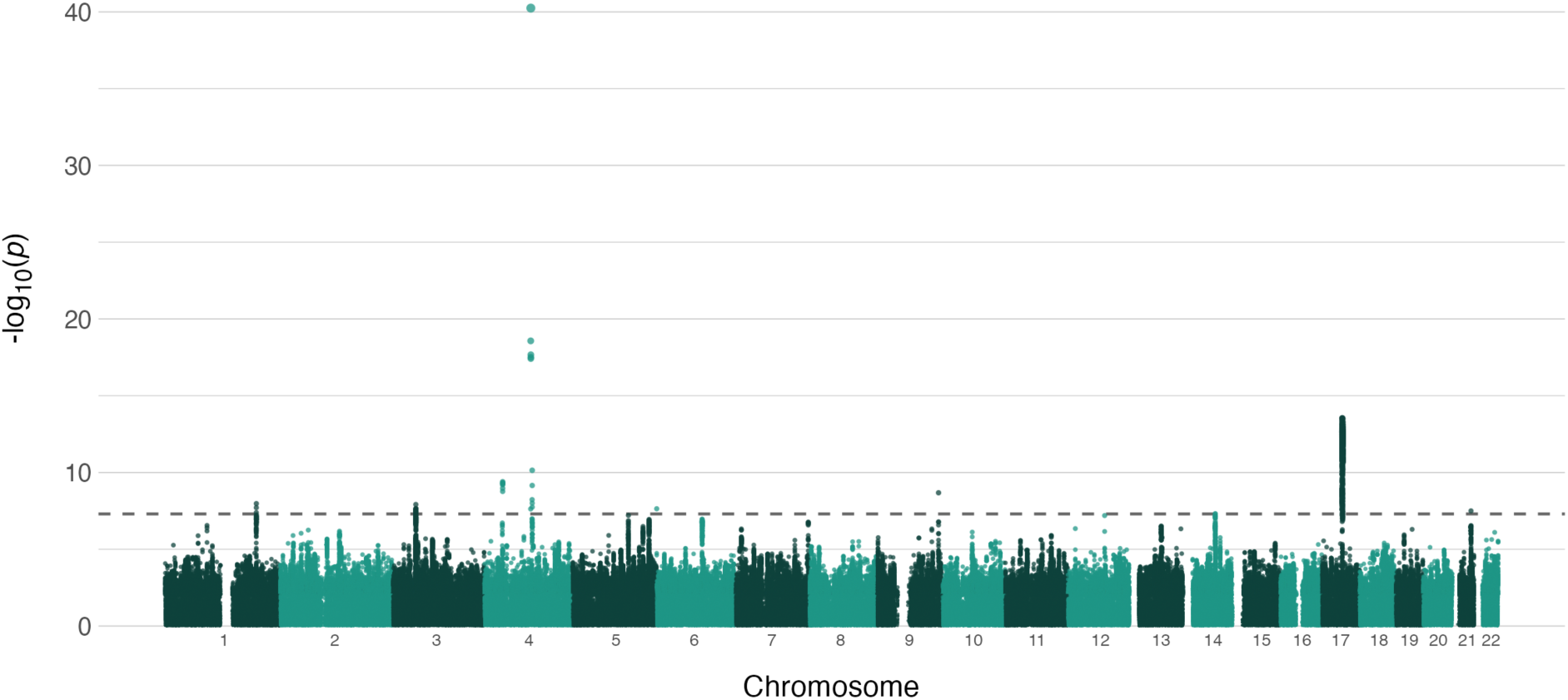
Manhattan plot of multivariate genome-wide association statistics for mental health. Manhattan plot showing the multivariate genome-wide association of our multivariate measure of mental health. We identified 10 loci associated with the multivariate genome-wide association statistics for mental health.

In order to identify shared genomic loci between the mental health profiles and psychiatric disorders, we used GWAS summary statistics from prior case-control studies including schizophrenia (SCZ)^1^, bipolar disorders (BIP)^4^, major depression (MD)^6^, attention-deficit hyperactivity disorder (ADHD)^32^, autism spectrum disorder (ASD)^33^, post-traumatic stress disorder (PTSD)^34^, and anxiety (ANX)^8^, see also Suppl. Table 2. First, we compared the gene set from the multivariate mental health genome-wide association statistics with the gene set from each of the psychiatric disorders. Here we found 35 overlapping genes, 29 with SCZ, 7 with BIP, and 1 with ADHD (see Suppl. Table 3). It is important to note that the 7 overlapping genes with BIP were mapped from only 2 separate loci. Next, we extracted the loci from each case-control GWAS (202 in total) and assessed whether each locus was significant in the multivariate genome-wide association statistics for mental health profiles as well. Of the 202 loci significant in any of the disorders, one showed genome-wide significance at P < 5e-8 and 122 showed nominal significance at P < 0.05 only in the multivariate genome-wide association statistics for mental health profiles, potentially indicating some shared but small effects.

Next, we explored the genetic overlap between the mental health profiles and the psychiatric disorders through the conjunctional false discovery rate (conjFDR)^31,35^ which leverages pleiotropy between two phenotypes to estimate shared genetic determinants. ConjFDR allows for the discovery of shared genetic determinants even when those loci are not genome-wide significant in either of the traits in the analysis. Through conjFDR we identified 35 overlapping loci in total between the multivariate genome-wide association statistics for mental health profiles and psychiatric disorders. We found 10 overlapping loci between the multivariate measure of mental health profiles and BIP, 8 overlapping loci with both MD and ADHD, 5 overlapping loci with SCZ, and 4 overlapping loci with autism (see Suppl. Figure 2). FUMA identified 89 genes associated with these loci (See Suppl. Table 3). We found no overlapping loci between the mental health profiles and ANX or PTSD, which may be related to the limited power in these GWASs (see Suppl. Table 2).

**Figure 2.**
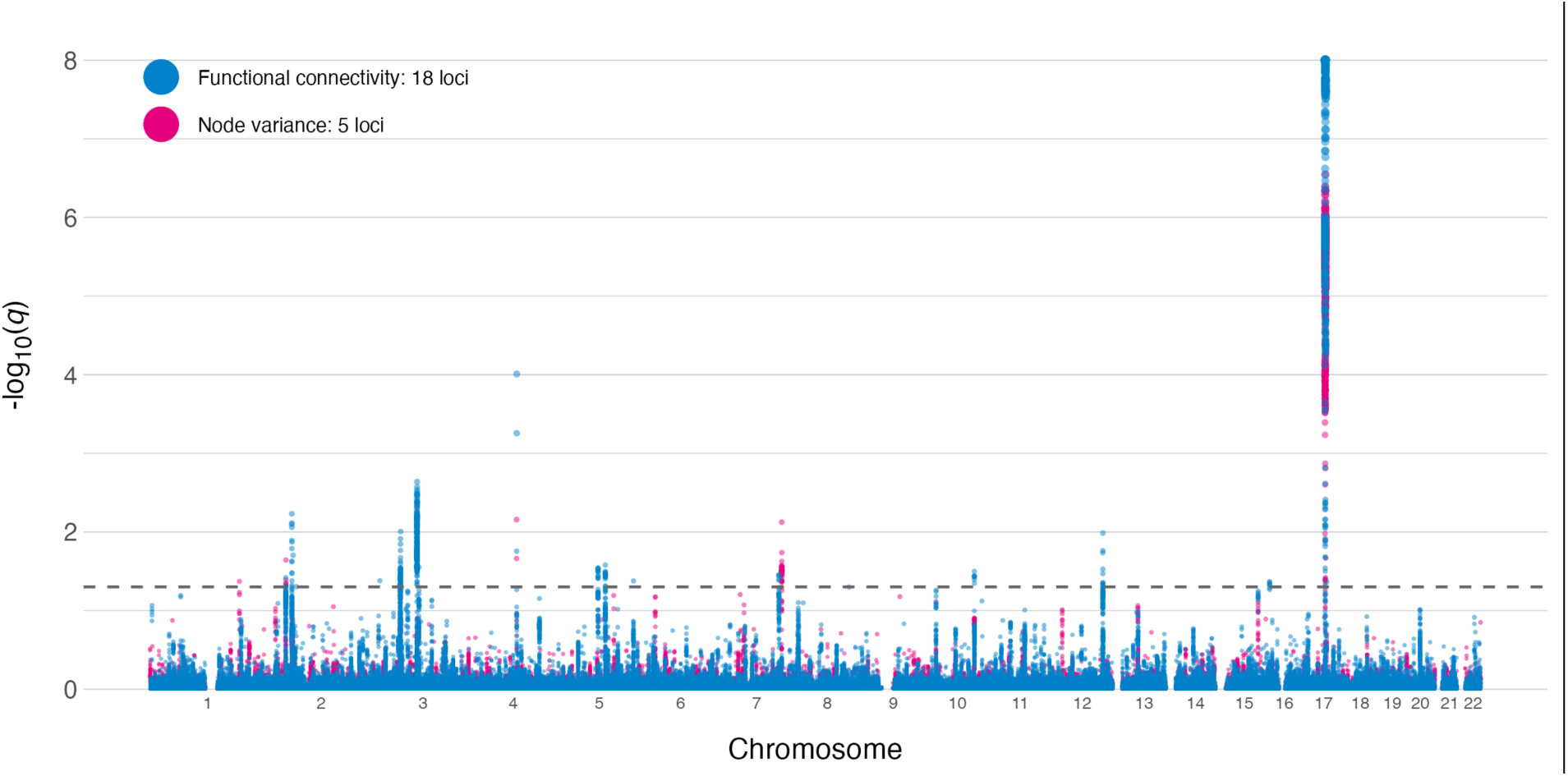
Manhattan plot of conjFDR between multivariate genome-wide association statistics for mental health and the brain functional connectome. Association strength per locus is depicted as q-value from the conjunctional FDR. Values for FC and node variance are shown in the same figure with separate colors.

We then calculated the number of shared genetic loci between the multivariate genome-wide association statistics for mental health profiles and the two multivariate measures of the brain functional connectome using conjFDR. We used the GWAS summary statistics from our previous study of brain function^13^. In contrast to the prior study in which we identified genetic overlap between brain function and psychiatric disorders, we here investigated overlaps with the multivariate genome-wide association statistics for mental health to investigate if this approach captures associations not revealed through case-control GWAS approaches. Figure 2 shows two Manhattan plots of the conjunctional FDR analyses between both functional connectivity and node variance with the multivariate GWAS on the multivariate genome-wide association statistics for mental health profiles. Genetic signal was adequate (See Suppl. Figure 3). The multivariate genome-wide association statistics for mental health profiles shared 18 loci with functional connectivity and 5 with node variance. A full list of genes associated with the (in total) 23 unique shared loci between the multivariate summary statistics and FC is presented in Suppl. Table 5. The number of overlapping loci between the brain functional connectome and the multivariate genome-wide association statistics for mental health profiles (18 for FC, 5 for node variance) was generally larger than the number of shared loci between the brain functional connectome and the psychiatric disorders (with the exception of SCZ) identified in our previous study^13^. When we mapped the genes from these loci using FUMA and tested for enrichment in gene-sets using the reactome toolbox^30^ we found that the genes associated with these shared loci are involved in a number of neurobiologically relevant processes such as axonal growth regulation (NGFR and RHOA) and regulation of transcription factors related through MECP2 (MEF2C, see Suppl. Table 6).

## Discussion

In this study we identified a number of loci associated with multivariate genome-wide association statistics for mental health profiles and found overlapping loci with the measures of brain function and psychiatric disorders. Using MOSTest we were able to leverage the phenotypic overlap between different mental health profiles to identify new loci associated with a multivariate measure of mental health. Genes associated with these loci showed regional expression in different parts of the brain (e.g. cerebellum, amygdala).

Our analysis using conjFDR revealed a number of shared loci and genes between the multivariate genome-wide association statistics and the psychiatric disorders. This demonstrates the shared genetics between psychiatric symptoms regardless of clinical diagnosis, emphasizes the utility of using population-level phenotypes to investigate variance in mental health profiles, and highlights the advantage in leveraging pleiotropy between complex phenotypes to boost discovery. We found shared genes with all but two case-control GWAS (ANX, PTSD), which also had the two smallest sample sizes, which may reflect insufficient power to detect an effect^36^, or may indicate the absence of an effect with those disorders. The largest overlap was with SCZ, which shared 29 genes in the geneset with the multivariate measure. Future sample increases in the case-control GWAS may reveal shared genetics with other complex traits, including population based mental health phenotypes and brain imaging features.

We also identified a number of overlapping loci between mental health profiles and fMRI measures of brain function, including 18 shared loci with functional connectivity and 5 shared loci with node variance. The higher number of shared loci for functional connectivity might be partially explained by the number of phenotypes in each composite measure. While the functional connectivity GWAS comprises 210 measures, i.e. partial correlations between 21 brain nodes, the node variance GWAS encompasses only the temporal variance in each node. It is possible that the number of phenotypes included in the multivariate analysis can affect the discovery^25^. Both the functional connectivity and node variance summary statistics had the same sample size (N=30.701). For our analyses this means that the difference in their overlap with the multivariate genome-wide association statistics for mental health is due to either the discrepancy in the number of features contained within the composite measure, or alternatively because of different biological processes underlying both measures. The measures differ in that functional connectivity refers to the correlation between brain networks (edge strength), which is possibly governed by different processes than the temporal variance in activity within brain networks. Overall, we found a number of genes associated with the shared loci that are involved in biologically relevant processes such as axonal growth and energy transport (See Suppl. Table 6). Although more thorough functional analysis is necessary, this could suggest that axonal growth processes is a shared feature between brain connectivity and mental disorders, which would be in line with previous evidence linking axonal growth with both processes independently^37–39^.

The two conjunctional analyses with fMRI measures and the multivariate genome-wide association statistics for mental health each showed a number of overlapping loci. Not all shared loci were unique, this can be partially explained by the definition of the brain networks in our analyses. The functional connectivity and the node variance measures use the same 21 nodes, and, ultimately, the two measures reflect different properties of the same time series. We found that the number of overlapping loci between the multivariate genome-wide association statistics for mental health and the brain functional connectome was generally larger than findings from our previous study highlighting shared genetic loci between psychiatric disorders and the brain functional connectome. This may partly be due to the larger sample size of the multivariate measure of mental health, but it could also reflect that the multivariate genome-wide association statistics capture genetic variance more generally related to the brain functional connectome. We found little direct overlap between loci of these two GWAS’ separately, which highlights the discovery boost advantage of using conjFDR in phenotypes with generally low heritability.

The main implication from our findings is that we can identify shared genetic variants between a multifactorial measure of mental health in an undiagnosed population sample and fMRI-based measures of brain functional connectivity. Several limitations should be considered. First, the data were obtained from a middle-aged and older White British population, which limits the generalizability of the findings. Further, the mental health questionnaires are self-administered, so the data is vulnerable to various response and self-selection biases^40^. We excluded individuals with a psychiatric diagnosis in our independent component analysis in order to maximalize the population variance and to mitigate the influence of a smaller number of individuals with a (diagnosed) psychiatric condition. This results in a healthier sample, lower variance on a number of severe symptom domains and possible survivor biases^24^. This bias also extends to the imaging data, where participants are reportedly healthier than the general population^41^. Further, MOSTest currently lacks effect direction. This complicates further analyses such as genetic correlations that require reliable effect directions. Nonetheless, FUMA and MAGMA revealed brain structures associated with these mapped genes, such as the cerebellum, amygdala, and various parts of the cortex, which have been linked to psychiatric disorders and symptoms. To what degree these shared genes can explain shared clinical characteristics such as symptoms is an important and relevant issue that needs to be answered in future studies.

In conclusion, our multivariate GWAS on 13 mental health symptom profiles showed a number of shared genetic loci with two fMRI-measures reflecting brain function and connectivity. This provides further genetic evidence of an association between brain function and mental health traits in the population.

## Data Availability

All data used in this study are part of the publicly available UK Biobank initiative (https://www.ukbiobank.ac.uk/). Summary statistics for the disorders are publicly available through their respective consortia. The summary statistics for the multivariate analyses will be shared on GitHub upon acceptance.

## Acknowledgements

The authors were funded by the Research Council of Norway (#276082 LifespanHealth, #223273 NORMENT, #283798 ERA-NET Neuron SYNSCHIZ, #249795), the South-East Norway Regional Health Authority (2019101, 2019107, and 2020086), and the European Research Council under the European Union’s Horizon2020 Research and Innovation program (ERC Starting Grant #802998), as well as the Horizon2020 Research and Innovation Action Grant CoMorMent (#847776). This research has been conducted using the UK Biobank Resource (access code 27412, https://www.ukbiobank.ac.uk/). E.T. has been supported by the Foundation “De Drie Lichten” and The Simons Foundation Fund in The Netherlands. This work was performed on the TSD (Tjenester for Sensitive Data) facilities, owned by the University of Oslo, operated and developed by the TSD service group at the University of Oslo, IT-Department (USIT). Computations were also performed on resources provided by UNINETT Sigma2 - the National Infrastructure for High Performance Computing and Data Storage in Norway.

## Conflicts of interest

D.R., D.A., O.F., D.vd.M., O.B.S., L.T.W. and T.K. declare no conflicts of interest. O.A.A. is a consultant to HealthLytix and received speakers honorarium from Lundbeck.

## Author contributions

D.R. and T.K. conceived the study; D.R. analyzed the data with contributions from T.K.; All authors contributed with conceptual input on methods and/or interpretation of results; D.R. and T.K. wrote the first draft of the paper and all authors contributed to the final manuscript.

## Code availability

Code will be made publicly available via GitHub (https://www.github.com/norment/open-science) upon acceptance of the manuscript.

## Supplementary Figures

**Suppl. Figure 1.**
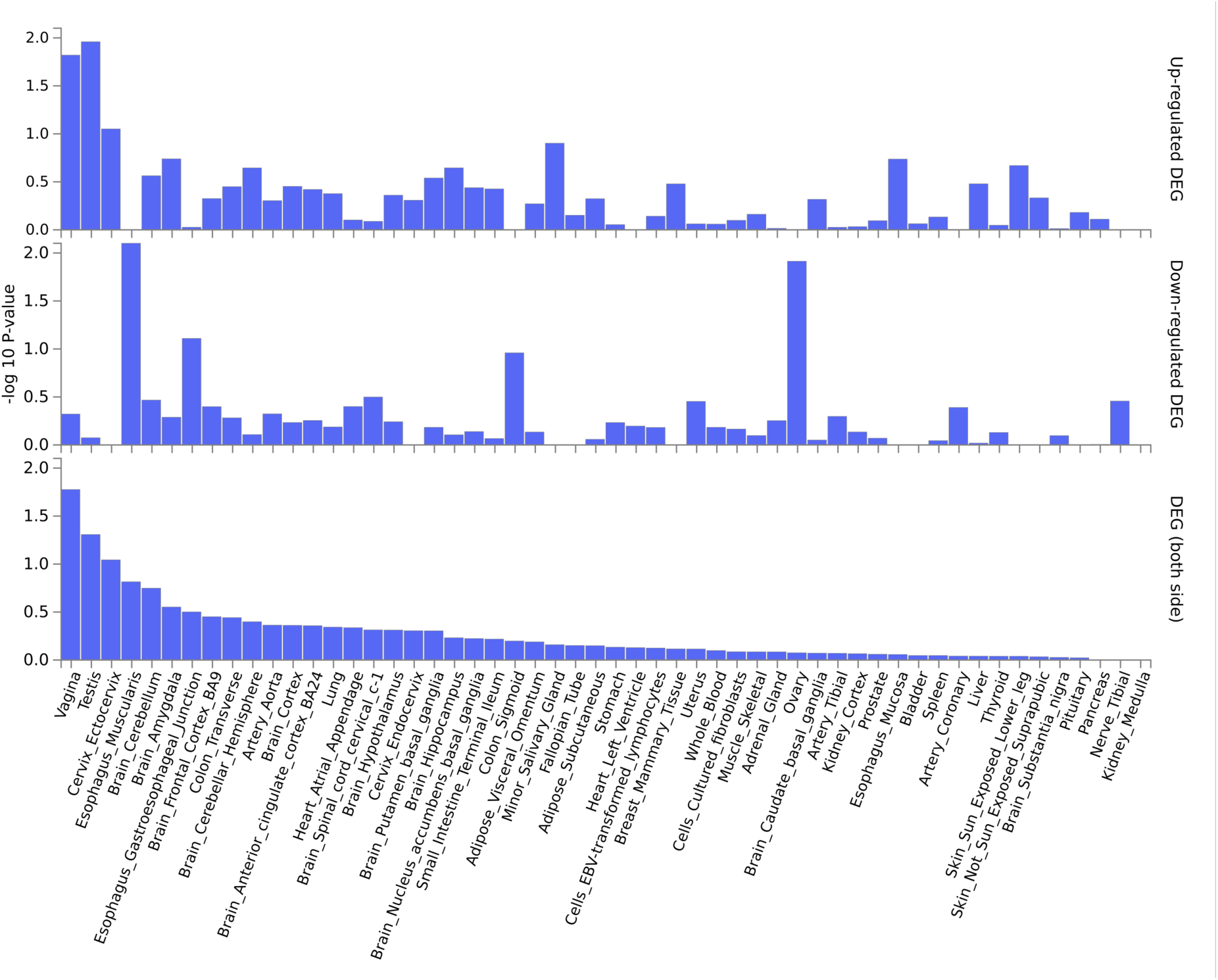
MAGMA output. List of biological structures mapped by MAGMA implemented through FUMA. Separate facets for up-regulated (top), down-regulated (middle), and both directions (bottom). Among some tangentially related structures (e.g. sexual organs, esophagus), there are large number of items on the higher end of the spectrum are related to brain structures (e.g. cerebellum, amygdala, cortex).

**Suppl. Figure 2.**
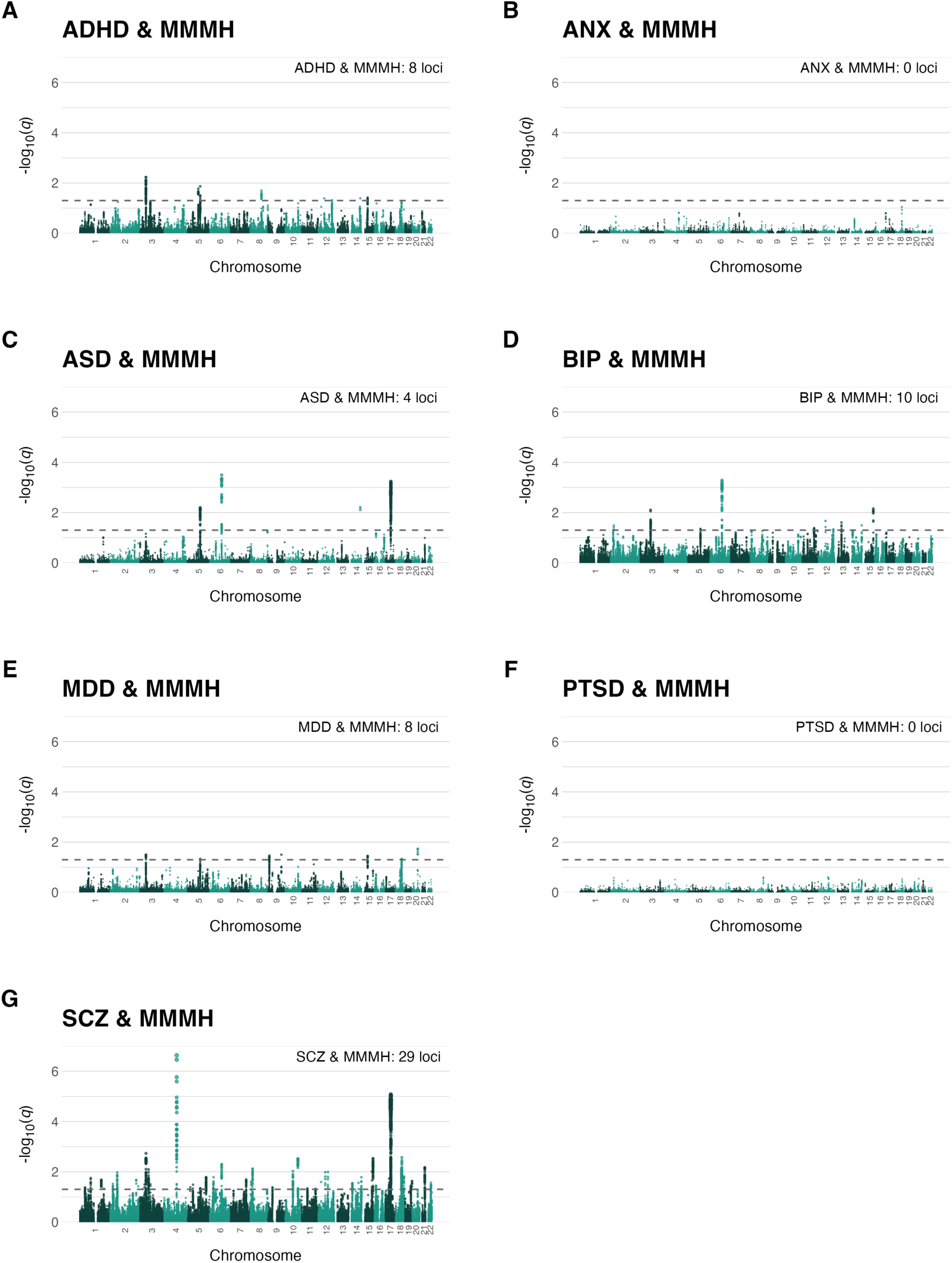
Manhattan plots from conjFDR between psychiatric disorders and the multivariate genome-wide association statistics for mental health. Manhattan plots illustrating the shared genetic determinants between a number of psychiatric disorders and the multivariate genome-wide association statistics for mental health. It shows the largest number of shared loci between the multivariate summary statistics and SCZ (29 loci), BIP (10 loci), followed by ADHD and MDD (both 8 loci).

**Suppl. Figure 3.**
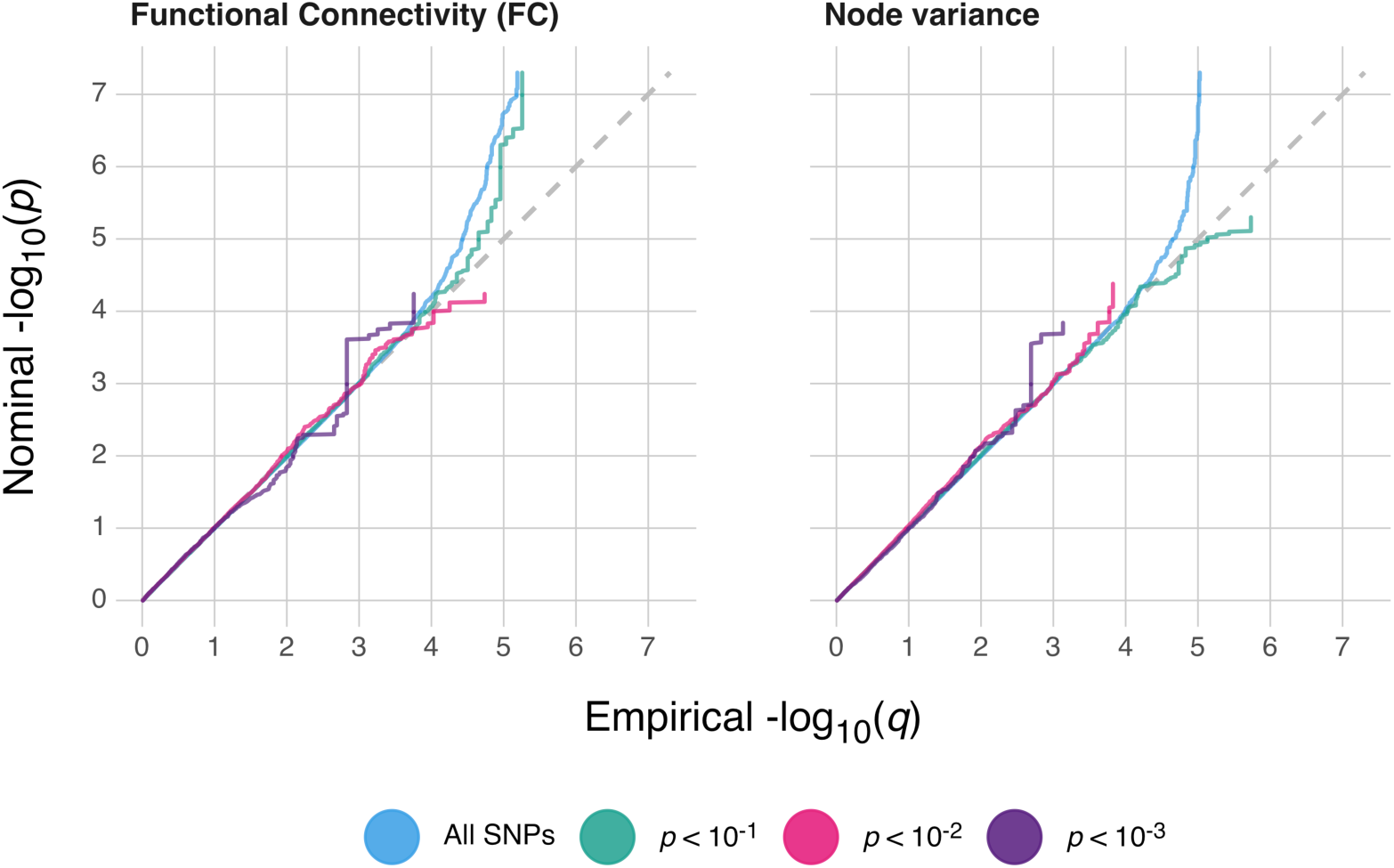
QQ plots of the genetic signal of the conjFDR between the multivariate genome-wide association statistics for mental health and the brain functional connectome. Figure showing no inflation of signal in the conjFDR at different thresholds. Genetic signal overall was not very strong, but sufficient for the analyses in this pipeline.

## Supplementary Tables

**Suppl. Table 1.**
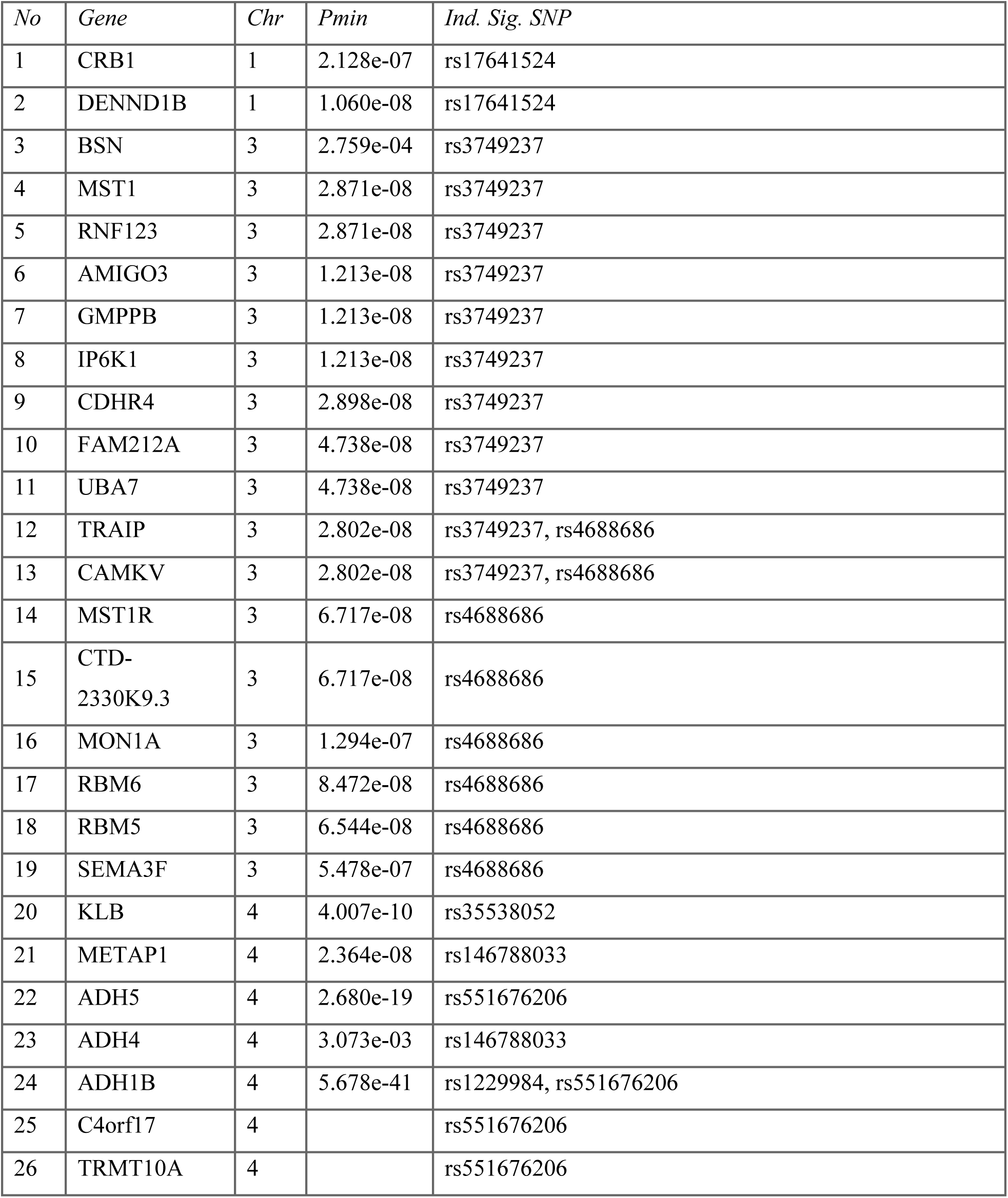
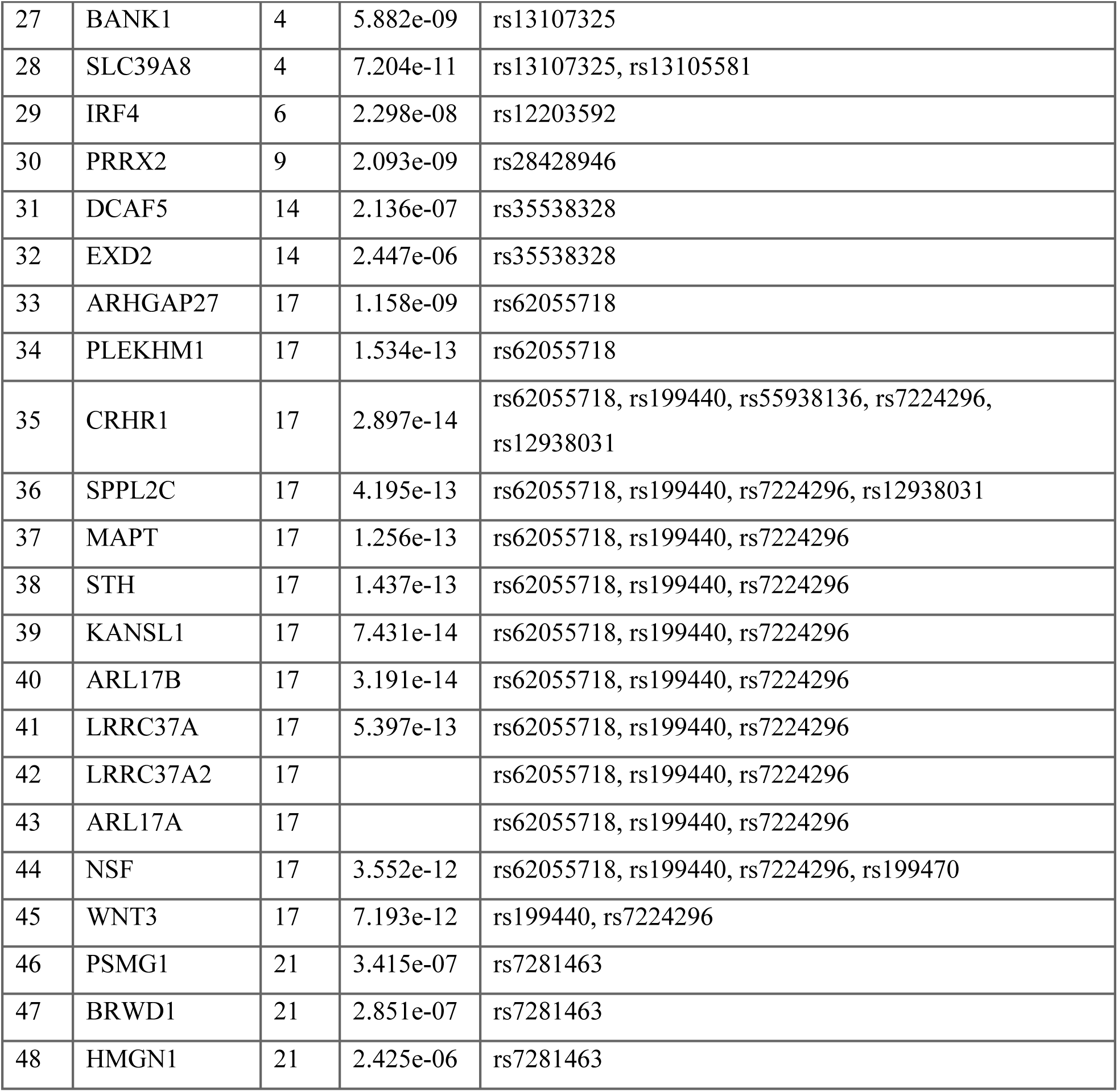
Mapped genes from the multivariate genome-wide association statistics for mental health. Genes mapped by FUMA on the summary statistics from the multivariate summary statistics. We found in total 48 genes associated with the genetic signal of the multivariate measure.

**Suppl. Table 2.** Overview of the genetic association analyses on psychiatric conditions used in this udy. List of GWASs we included in the genetic analyses in this paper and their meta data. Sample size in e table and in the summary statistics used in the final analyses may differ since we excluded individuals ready included in the UK Biobank and included only individuals with White European ancestry.

| **Suppl. Table 2. Overview of the genetic association analyses on psychiatric conditions used in this study** List of GWASs we included in the genetic analyses in this paper and their meta data. Sample size in the table and in the summary statistics used in the final analyses may differ since we excluded individuals already included in the UK Biobank and included only individuals with White European ancestry. |  |  |  |  |  |  |  |  |
| --- | --- | --- | --- | --- | --- | --- | --- | --- |
| *Pheno-type* | *Consortium* | *Sample* | *Citation* | *N<sub>case</sub>* | *N<sub>control</sub>* | *Number of loci* | *Lambda<sub>GC</sub>* | *h<sup>2</sup> (se)* |
| SCZ | PGC | Meta analysis of European individuals in PGC and CLOZUK samples | Pardiñas et al., 2018 | 40,675 | 64,643 | 135 | 1.6831 | 0.41 (0.01) |
| BIP | PGC | European individuals | Mullins et al., 2021 | 41,917 | 371,549 | 44 | 1.4316 | 0.0853 (<0.01) |
| MDD | PGC | European individuals | Wray et al., 2018 | 69,576 | 161,613 | 2 | 1.1973 | 0.0717 (<0.01) |
| ADHD | PGC | European individuals | Demontis et al., 2019 | 19,099 | 34,194 | 12 | 1.2531 | 0.2354 (0.02) |
| ASD | PGC, iPSYCH | Meta analysis of European individuals | Grove et al., 2019 | 18,381 | 27,969 | 2 | 1.1715 | 0.1941 (0.02) |
| PTSD | PGC | European individuals | Duncan et al., 2018 | 2,424 | 7,113 | 0 | 1.0165 | 0.101 (0.04) |
| ANX | ANGST | Meta analysis of European individuals | Otowa et al., 2016 | 7,016 | 14,745 | 1 | 1.0345 | 0.0763 (0.03) |

**Suppl. Table 3.** Number of overlapping genes in gene sets with the multivariate genome-wide association statistics for mental health (MMMH) Table showing the number of overlapping genes identified through either overlap in gene sets, the number of genes from the disorder GWAS alone, and the number of shared genes identified through conjFDR.

| *Disorder* | *Measure* | *Number of overlapping genes in gene sets* | *Number of genes in disorder GWAS alone* | *Number of shared genes identified through cFDR* |
| --- | --- | --- | --- | --- |
| ADHD | MMMH | 1 | 22 | 37 |
| ANX | MMMH | 0 | 0 | 0 |
| ASD | MMMH | 0 | 6 | 16 |
| BIP | MMMH | 7 | 184 | 28 |
| MDD | MMMH | 0 | 1 | 29 |
| PTSD | MMMH | 0 | 0 | 0 |
| SCZ | MMMH | 27 | 600 | 72 |

**Suppl. Table 4.**
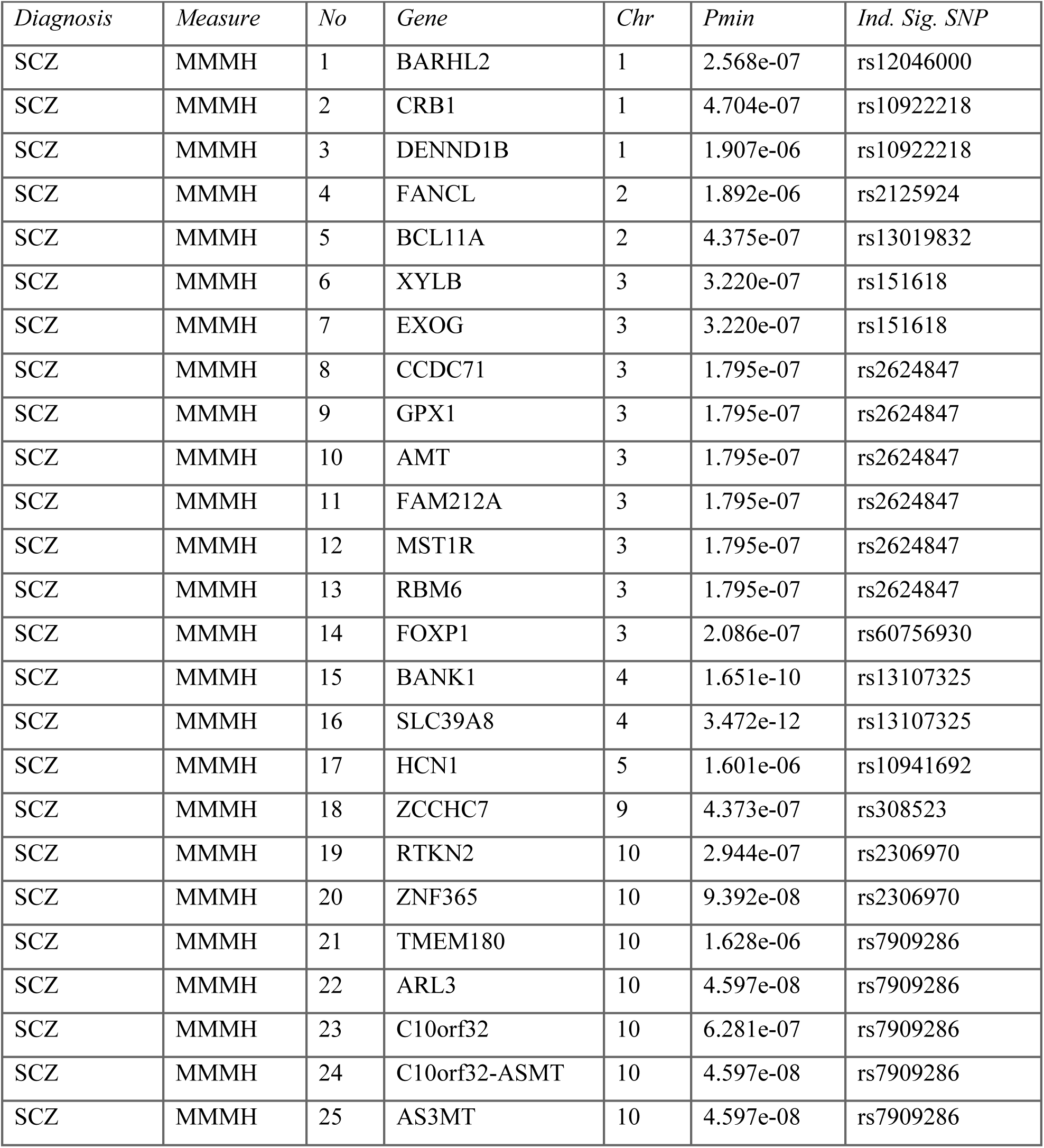
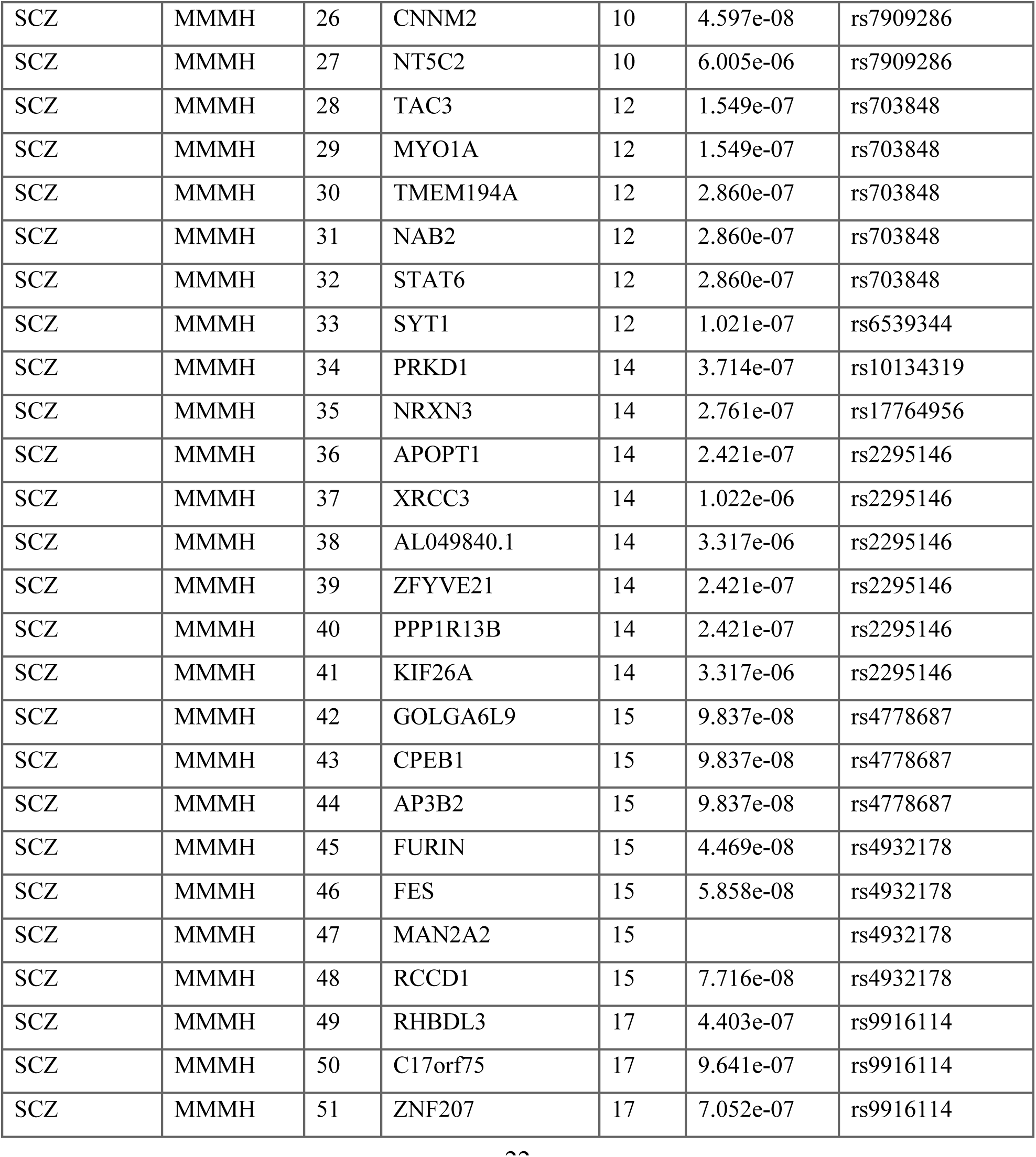
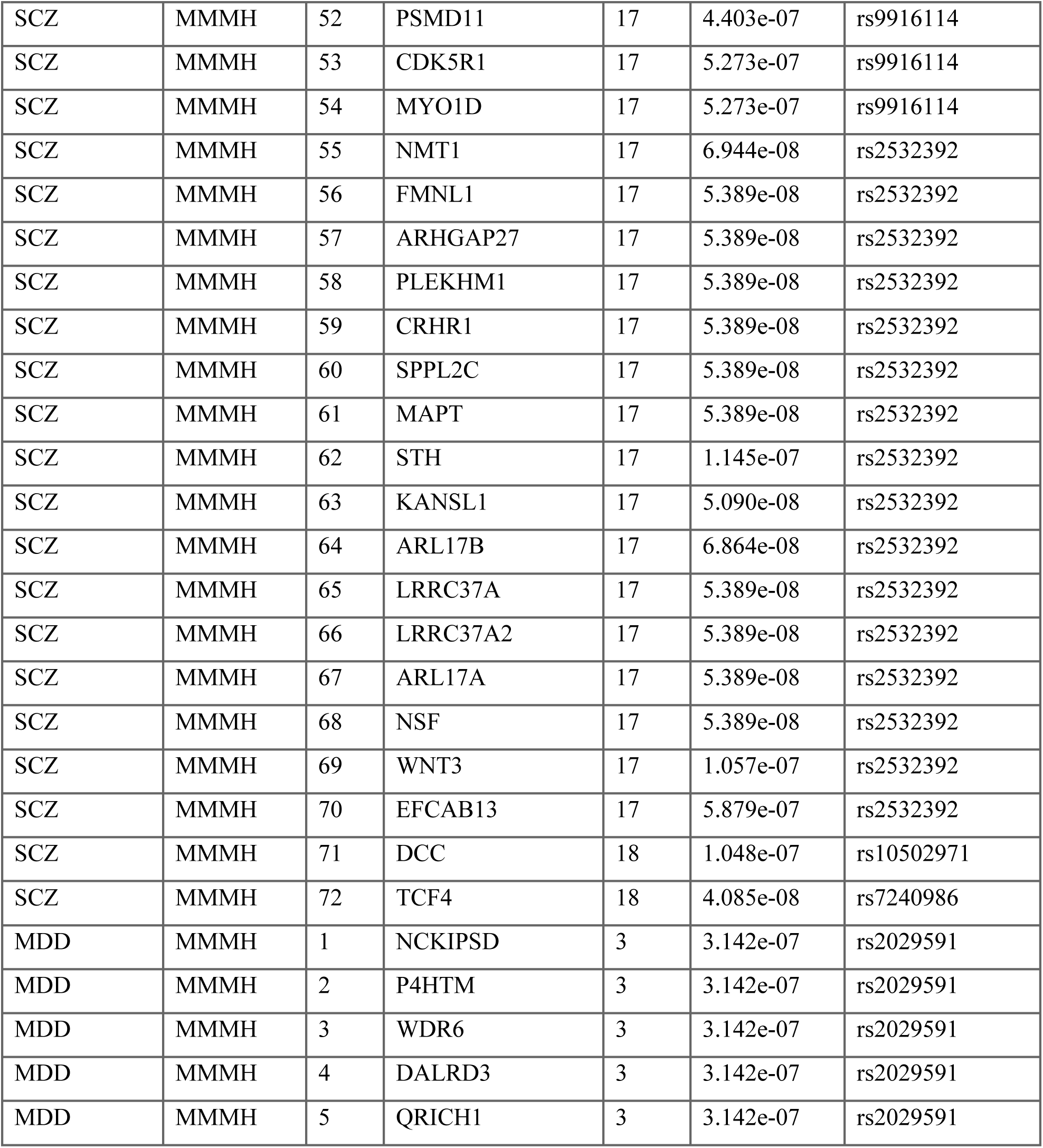
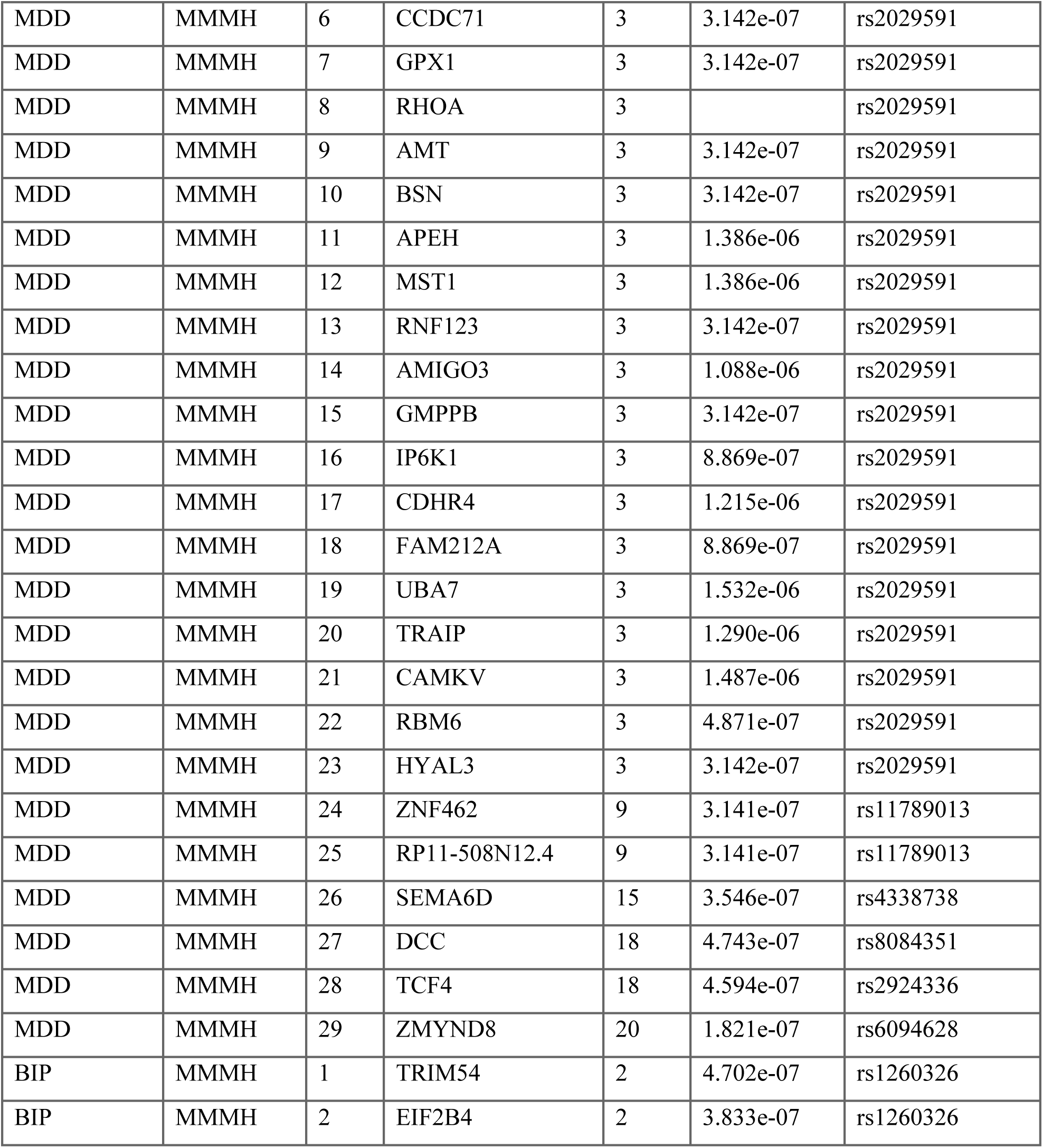
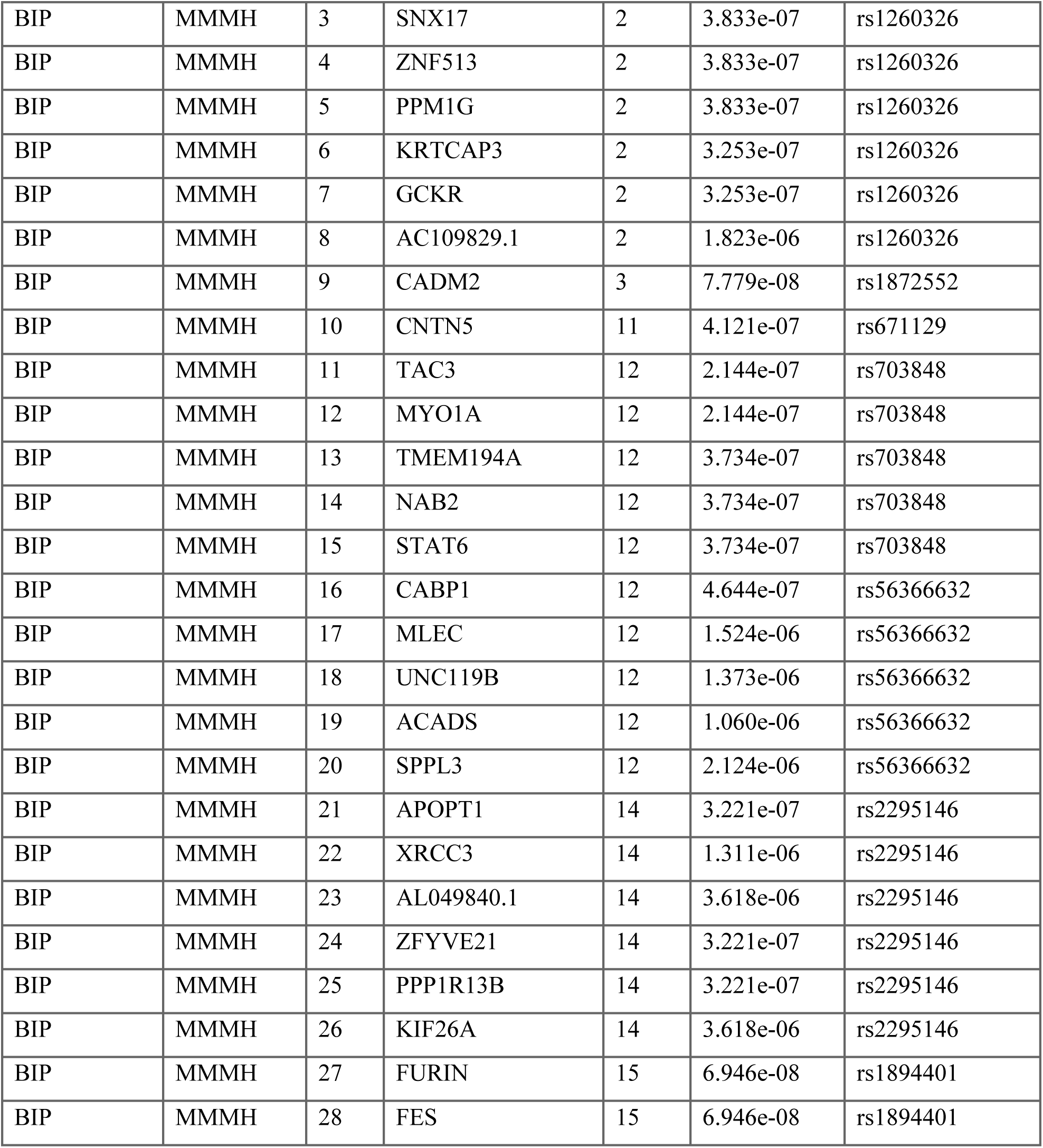
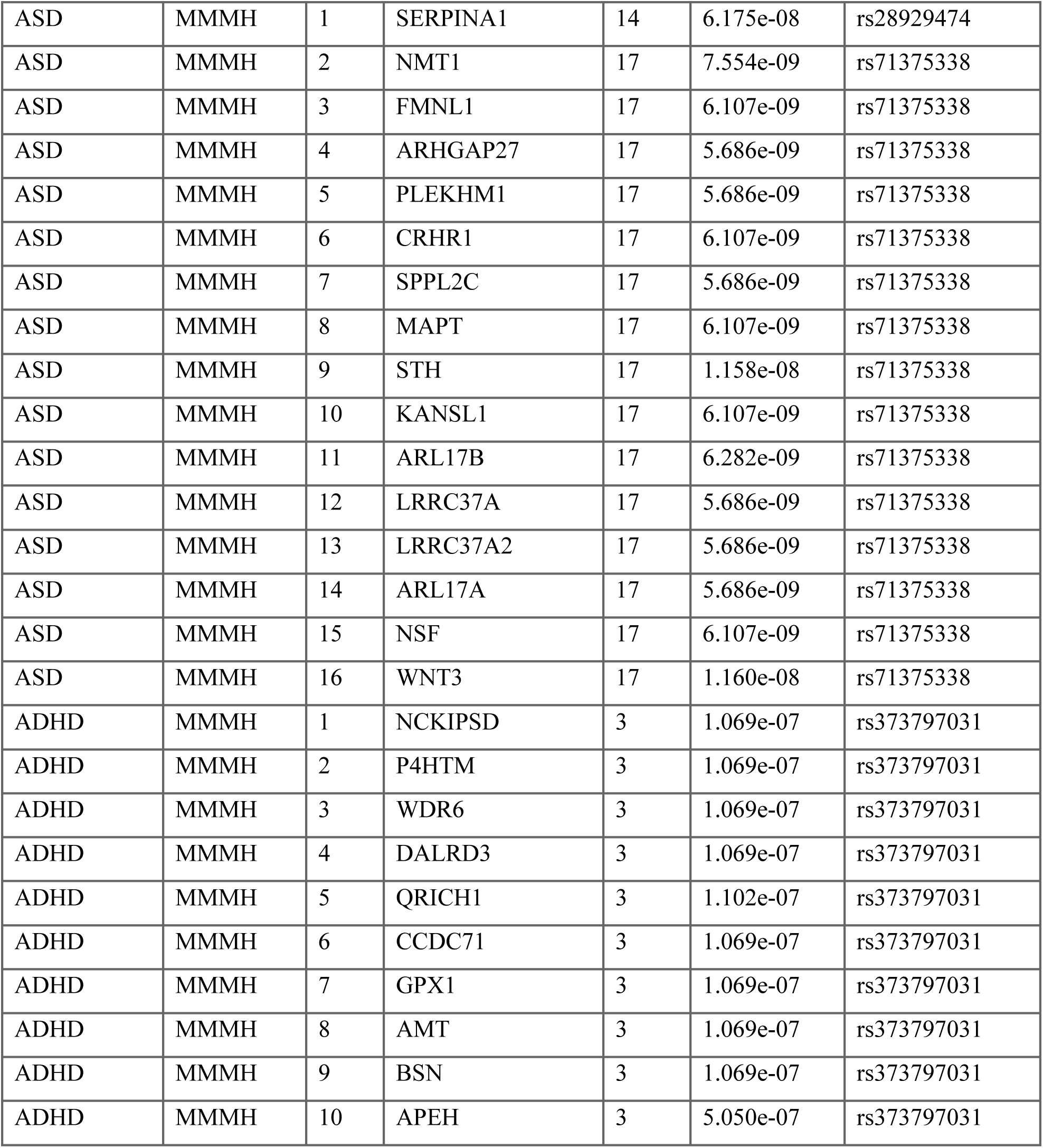
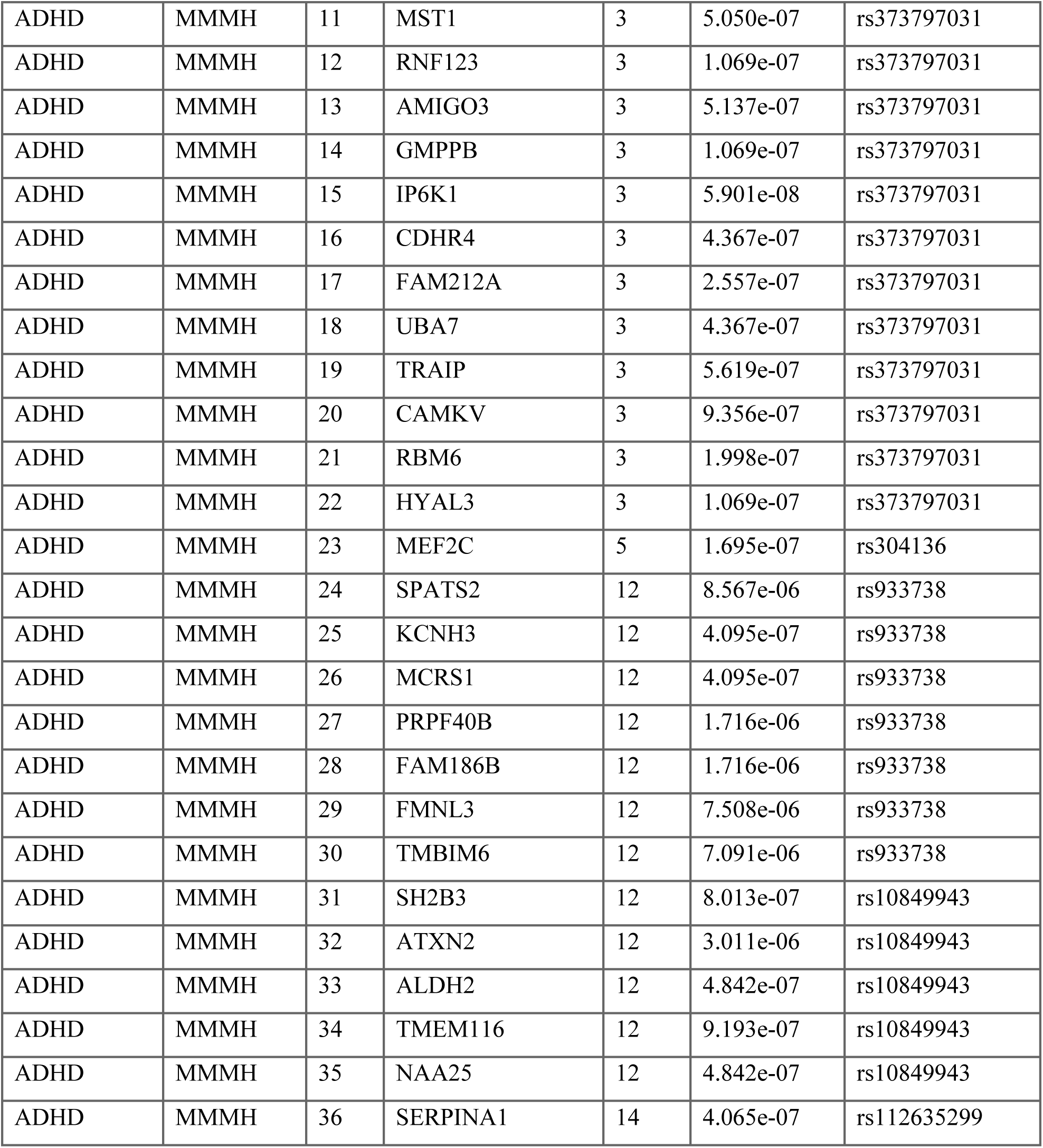
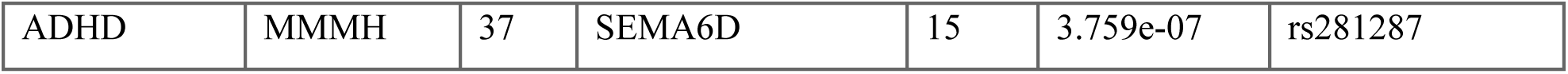
List of genes associated with the shared genetic determinants between the psychiatric disorders and multivariate genome-wide association statistics for mental health. Genes associated with conjFDR output from diagnosis and multivariate genome-wide association statistics for mental health. This list only includes psychiatric disorders for which we identified significant loci. The number of mapped genes can be greater than the number of discovered loci since loci can be associated with more than one gene.

| *Diagnosis* | *Measure* | *No* | *Gene* | *Chr* | *Pmin* | *Ind. Sig. SNP* |
| --- | --- | --- | --- | --- | --- | --- |
| SCZ | MMMH | 1 | BARHL2 | 1 | 2.568e-07 | rs12046000 |
| SCZ | MMMH | 2 | CRB1 | 1 | 4.704e-07 | rs10922218 |
| SCZ | MMMH | 3 | DENND1B | 1 | 1.907e-06 | rs10922218 |
| SCZ | MMMH | 4 | FANCL | 2 | 1.892e-06 | rs2125924 |
| SCZ | MMMH | 5 | BCL11A | 2 | 4.375e-07 | rs13019832 |
| SCZ | MMMH | 6 | XYLB | 3 | 3.220e-07 | rs151618 |
| SCZ | MMMH | 7 | EXOG | 3 | 3.220e-07 | rs151618 |
| SCZ | MMMH | 8 | CCDC71 | 3 | 1.795e-07 | rs2624847 |
| SCZ | MMMH | 9 | GPX1 | 3 | 1.795e-07 | rs2624847 |
| SCZ | MMMH | 10 | AMT | 3 | 1.795e-07 | rs2624847 |
| SCZ | MMMH | 11 | FAM212A | 3 | 1.795e-07 | rs2624847 |
| SCZ | MMMH | 12 | MST1R | 3 | 1.795e-07 | rs2624847 |
| SCZ | MMMH | 13 | RBM6 | 3 | 1.795e-07 | rs2624847 |
| SCZ | MMMH | 14 | FOXP1 | 3 | 2.086e-07 | rs60756930 |
| SCZ | MMMH | 15 | BANK1 | 4 | 1.651e-10 | rs13107325 |
| SCZ | MMMH | 16 | SLC39A8 | 4 | 3.472e-12 | rs13107325 |
| SCZ | MMMH | 17 | HCN1 | 5 | 1.601e-06 | rs10941692 |
| SCZ | MMMH | 18 | ZCCHC7 | 9 | 4.373e-07 | rs308523 |
| SCZ | MMMH | 19 | RTKN2 | 10 | 2.944e-07 | rs2306970 |
| SCZ | MMMH | 20 | ZNF365 | 10 | 9.392e-08 | rs2306970 |
| SCZ | MMMH | 21 | TMEM180 | 10 | 1.628e-06 | rs7909286 |
| SCZ | MMMH | 22 | ARL3 | 10 | 4.597e-08 | rs7909286 |
| SCZ | MMMH | 23 | C10orf32 | 10 | 6.281e-07 | rs7909286 |
| SCZ | MMMH | 24 | C10orf32-ASMT | 10 | 4.597e-08 | rs7909286 |
| SCZ | MMMH | 25 | AS3MT | 10 | 4.597e-08 | rs7909286 |

|  |  |  |  |  |  |  |
| --- | --- | --- | --- | --- | --- | --- |
| *Diagnosis* | *Measure* | *No* | *Gene* | *Chr* | *Pmin* | *Ind. Sig. SNP* |
| SCZ | MMMH | 26 | CNNM2 | 10 | 4.597e-08 | rs7909286 |
| SCZ | MMMH | 27 | NT5C2 | 10 | 6.005e-06 | rs7909286 |
| SCZ | MMMH | 28 | TAC3 | 12 | 1.549e-07 | rs703848 |
| SCZ | MMMH | 29 | MYO1A | 12 | 1.549e-07 | rs703848 |
| SCZ | MMMH | 30 | TMEM194A | 12 | 2.860e-07 | rs703848 |
| SCZ | MMMH | 31 | NAB2 | 12 | 2.860e-07 | rs703848 |
| SCZ | MMMH | 32 | STAT6 | 12 | 2.860e-07 | rs703848 |
| SCZ | MMMH | 33 | SYT1 | 12 | 1.021e-07 | rs6539344 |
| SCZ | MMMH | 34 | PRKD1 | 14 | 3.714e-07 | rs10134319 |
| SCZ | MMMH | 35 | NRXN3 | 14 | 2.761e-07 | rs17764956 |
| SCZ | MMMH | 36 | APOPT1 | 14 | 2.421e-07 | rs2295146 |
| SCZ | MMMH | 37 | XRCC3 | 14 | 1.022e-06 | rs2295146 |
| SCZ | MMMH | 38 | AL049840.1 | 14 | 3.317e-06 | rs2295146 |
| SCZ | MMMH | 39 | ZFYVE21 | 14 | 2.421e-07 | rs2295146 |
| SCZ | MMMH | 40 | PPP1R13B | 14 | 2.421e-07 | rs2295146 |
| SCZ | MMMH | 41 | KIF26A | 14 | 3.317e-06 | rs2295146 |
| SCZ | MMMH | 42 | GOLGA6L9 | 15 | 9.837e-08 | rs4778687 |
| SCZ | MMMH | 43 | CPEB1 | 15 | 9.837e-08 | rs4778687 |
| SCZ | MMMH | 44 | AP3B2 | 15 | 9.837e-08 | rs4778687 |
| SCZ | MMMH | 45 | FURIN | 15 | 4.469e-08 | rs4932178 |
| SCZ | MMMH | 46 | FES | 15 | 5.858e-08 | rs4932178 |
| SCZ | MMMH | 47 | MAN2A2 | 15 |  | rs4932178 |
| SCZ | MMMH | 48 | RCCD1 | 15 | 7.716e-08 | rs4932178 |
| SCZ | MMMH | 49 | RHBDL3 | 17 | 4.403e-07 | rs9916114 |
| SCZ | MMMH | 50 | C17orf75 | 17 | 9.641e-07 | rs9916114 |
| SCZ | MMMH | 51 | ZNF207 | 17 | 7.052e-07 | rs9916114 |
| *Diagnosis* | *Measure* | *No* | *Gene* | *Chr* | *Pmin* | *Ind. Sig. SNP* |
| SCZ | MMMH | 52 | PSMD11 | 17 | 4.403e-07 | rs9916114 |
| SCZ | MMMH | 53 | CDK5R1 | 17 | 5.273e-07 | rs9916114 |
| SCZ | MMMH | 54 | MYO1D | 17 | 5.273e-07 | rs9916114 |
| SCZ | MMMH | 55 | NMT1 | 17 | 6.944e-08 | rs2532392 |
| SCZ | MMMH | 56 | FMNL1 | 17 | 5.389e-08 | rs2532392 |
| SCZ | MMMH | 57 | ARHGAP27 | 17 | 5.389e-08 | rs2532392 |
| SCZ | MMMH | 58 | PLEKHM1 | 17 | 5.389e-08 | rs2532392 |
| SCZ | MMMH | 59 | CRHR1 | 17 | 5.389e-08 | rs2532392 |
| SCZ | MMMH | 60 | SPPL2C | 17 | 5.389e-08 | rs2532392 |
| SCZ | MMMH | 61 | MAPT | 17 | 5.389e-08 | rs2532392 |
| SCZ | MMMH | 62 | STH | 17 | 1.145e-07 | rs2532392 |
| SCZ | MMMH | 63 | KANSL1 | 17 | 5.090e-08 | rs2532392 |
| SCZ | MMMH | 64 | ARL17B | 17 | 6.864e-08 | rs2532392 |
| SCZ | MMMH | 65 | LRRC37A | 17 | 5.389e-08 | rs2532392 |
| SCZ | MMMH | 66 | LRRC37A2 | 17 | 5.389e-08 | rs2532392 |
| SCZ | MMMH | 67 | ARL17A | 17 | 5.389e-08 | rs2532392 |
| SCZ | MMMH | 68 | NSF | 17 | 5.389e-08 | rs2532392 |
| SCZ | MMMH | 69 | WNT3 | 17 | 1.057e-07 | rs2532392 |
| SCZ | MMMH | 70 | EFCAB13 | 17 | 5.879e-07 | rs2532392 |
| SCZ | MMMH | 71 | DCC | 18 | 1.048e-07 | rs10502971 |
| SCZ | MMMH | 72 | TCF4 | 18 | 4.085e-08 | rs7240986 |
| MDD | MMMH | 1 | NCKIPSD | 3 | 3.142e-07 | rs2029591 |
| MDD | MMMH | 2 | P4HTM | 3 | 3.142e-07 | rs2029591 |
| MDD | MMMH | 3 | WDR6 | 3 | 3.142e-07 | rs2029591 |
| MDD | MMMH | 4 | DALRD3 | 3 | 3.142e-07 | rs2029591 |
| MDD | MMMH | 5 | QRICH1 | 3 | 3.142e-07 | rs2029591 |
| *Diagnosis* | *Measure* | *No* | *Gene* | *Chr* | *Pmin* | *Ind. Sig. SNP* |
| MDD | MMMH | 6 | CCDC71 | 3 | 3.142e-07 | rs2029591 |
| MDD | MMMH | 7 | GPX1 | 3 | 3.142e-07 | rs2029591 |
| MDD | MMMH | 8 | RHOA | 3 |  | rs2029591 |
| MDD | MMMH | 9 | AMT | 3 | 3.142e-07 | rs2029591 |
| MDD | MMMH | 10 | BSN | 3 | 3.142e-07 | rs2029591 |
| MDD | MMMH | 11 | APEH | 3 | 1.386e-06 | rs2029591 |
| MDD | MMMH | 12 | MST1 | 3 | 1.386e-06 | rs2029591 |
| MDD | MMMH | 13 | RNF123 | 3 | 3.142e-07 | rs2029591 |
| MDD | MMMH | 14 | AMIGO3 | 3 | 1.088e-06 | rs2029591 |
| MDD | MMMH | 15 | GMPPB | 3 | 3.142e-07 | rs2029591 |
| MDD | MMMH | 16 | IP6K1 | 3 | 8.869e-07 | rs2029591 |
| MDD | MMMH | 17 | CDHR4 | 3 | 1.215e-06 | rs2029591 |
| MDD | MMMH | 18 | FAM212A | 3 | 8.869e-07 | rs2029591 |
| MDD | MMMH | 19 | UBA7 | 3 | 1.532e-06 | rs2029591 |
| MDD | MMMH | 20 | TRAIP | 3 | 1.290e-06 | rs2029591 |
| MDD | MMMH | 21 | CAMKV | 3 | 1.487e-06 | rs2029591 |
| MDD | MMMH | 22 | RBM6 | 3 | 4.871e-07 | rs2029591 |
| MDD | MMMH | 23 | HYAL3 | 3 | 3.142e-07 | rs2029591 |
| MDD | MMMH | 24 | ZNF462 | 9 | 3.141e-07 | rs11789013 |
| MDD | MMMH | 25 | RP11-508N12.4 | 9 | 3.141e-07 | rs11789013 |
| MDD | MMMH | 26 | SEMA6D | 15 | 3.546e-07 | rs4338738 |
| MDD | MMMH | 27 | DCC | 18 | 4.743e-07 | rs8084351 |
| MDD | MMMH | 28 | TCF4 | 18 | 4.594e-07 | rs2924336 |
| MDD | MMMH | 29 | ZMYND8 | 20 | 1.821e-07 | rs6094628 |
| BIP | MMMH | 1 | TRIM54 | 2 | 4.702e-07 | rs1260326 |
| BIP | MMMH | 2 | EIF2B4 | 2 | 3.833e-07 | rs1260326 |
| BIP | MMMH | 3 | SNX17 | 2 | 3.833e-07 | rs1260326 |
| BIP | MMMH | 4 | ZNF513 | 2 | 3.833e-07 | rs1260326 |
| BIP | MMMH | 5 | PPM1G | 2 | 3.833e-07 | rs1260326 |
| BIP | MMMH | 6 | KRTCAP3 | 2 | 3.253e-07 | rs1260326 |
| BIP | MMMH | 7 | GCKR | 2 | 3.253e-07 | rs1260326 |
| BIP | MMMH | 8 | AC109829.1 | 2 | 1.823e-06 | rs1260326 |
| BIP | MMMH | 9 | CADM2 | 3 | 7.779e-08 | rs1872552 |
| BIP | MMMH | 10 | CNTN5 | 11 | 4.121e-07 | rs671129 |
| BIP | MMMH | 11 | TAC3 | 12 | 2.144e-07 | rs703848 |
| BIP | MMMH | 12 | MYO1A | 12 | 2.144e-07 | rs703848 |
| BIP | MMMH | 13 | TMEM194A | 12 | 3.734e-07 | rs703848 |
| BIP | MMMH | 14 | NAB2 | 12 | 3.734e-07 | rs703848 |
| BIP | MMMH | 15 | STAT6 | 12 | 3.734e-07 | rs703848 |
| BIP | MMMH | 16 | CABP1 | 12 | 4.644e-07 | rs56366632 |
| BIP | MMMH | 17 | MLEC | 12 | 1.524e-06 | rs56366632 |
| BIP | MMMH | 18 | UNC119B | 12 | 1.373e-06 | rs56366632 |
| BIP | MMMH | 19 | ACADS | 12 | 1.060e-06 | rs56366632 |
| BIP | MMMH | 20 | SPPL3 | 12 | 2.124e-06 | rs56366632 |
| BIP | MMMH | 21 | APOPT1 | 14 | 3.221e-07 | rs2295146 |
| BIP | MMMH | 22 | XRCC3 | 14 | 1.311e-06 | rs2295146 |
| BIP | MMMH | 23 | AL049840.1 | 14 | 3.618e-06 | rs2295146 |
| BIP | MMMH | 24 | ZFYVE21 | 14 | 3.221e-07 | rs2295146 |
| BIP | MMMH | 25 | PPP1R13B | 14 | 3.221e-07 | rs2295146 |
| BIP | MMMH | 26 | KIF26A | 14 | 3.618e-06 | rs2295146 |
| BIP | MMMH | 27 | FURIN | 15 | 6.946e-08 | rs1894401 |
| BIP | MMMH | 28 | FES | 15 | 6.946e-08 | rs1894401 |

|  |  |  |  |  |  |  |
| --- | --- | --- | --- | --- | --- | --- |
| *Diagnosis* | *Measure* | *No* | *Gene* | *Chr* | *Pmin* | *Ind. Sig. SNP* |
| ASD | MMMH | 1 | SERPINA1 | 14 | 6.175e-08 | rs28929474 |
| ASD | MMMH | 2 | NMT1 | 17 | 7.554e-09 | rs71375338 |
| ASD | MMMH | 3 | FMNL1 | 17 | 6.107e-09 | rs71375338 |
| ASD | MMMH | 4 | ARHGAP27 | 17 | 5.686e-09 | rs71375338 |
| ASD | MMMH | 5 | PLEKHM1 | 17 | 5.686e-09 | rs71375338 |
| ASD | MMMH | 6 | CRHR1 | 17 | 6.107e-09 | rs71375338 |
| ASD | MMMH | 7 | SPPL2C | 17 | 5.686e-09 | rs71375338 |
| ASD | MMMH | 8 | MAPT | 17 | 6.107e-09 | rs71375338 |
| ASD | MMMH | 9 | STH | 17 | 1.158e-08 | rs71375338 |
| ASD | MMMH | 10 | KANSL1 | 17 | 6.107e-09 | rs71375338 |
| ASD | MMMH | 11 | ARL17B | 17 | 6.282e-09 | rs71375338 |
| ASD | MMMH | 12 | LRRC37A | 17 | 5.686e-09 | rs71375338 |
| ASD | MMMH | 13 | LRRC37A2 | 17 | 5.686e-09 | rs71375338 |
| ASD | MMMH | 14 | ARL17A | 17 | 5.686e-09 | rs71375338 |
| ASD | MMMH | 15 | NSF | 17 | 6.107e-09 | rs71375338 |
| ASD | MMMH | 16 | WNT3 | 17 | 1.160e-08 | rs71375338 |
| ADHD | MMMH | 1 | NCKIPSD | 3 | 1.069e-07 | rs373797031 |
| ADHD | MMMH | 2 | P4HTM | 3 | 1.069e-07 | rs373797031 |
| ADHD | MMMH | 3 | WDR6 | 3 | 1.069e-07 | rs373797031 |
| ADHD | MMMH | 4 | DALRD3 | 3 | 1.069e-07 | rs373797031 |
| ADHD | MMMH | 5 | QRICH1 | 3 | 1.102e-07 | rs373797031 |
| ADHD | MMMH | 6 | CCDC71 | 3 | 1.069e-07 | rs373797031 |
| ADHD | MMMH | 7 | GPX1 | 3 | 1.069e-07 | rs373797031 |
| ADHD | MMMH | 8 | AMT | 3 | 1.069e-07 | rs373797031 |
| ADHD | MMMH | 9 | BSN | 3 | 1.069e-07 | rs373797031 |
| ADHD | MMMH | 10 | APEH | 3 | 5.050e-07 | rs373797031 |
| ADHD | MMMH | 11 | MST1 | 3 | 5.050e-07 | rs373797031 |
| ADHD | MMMH | 12 | RNF123 | 3 | 1.069e-07 | rs373797031 |
| ADHD | MMMH | 13 | AMIGO3 | 3 | 5.137e-07 | rs373797031 |
| ADHD | MMMH | 14 | GMPPB | 3 | 1.069e-07 | rs373797031 |
| ADHD | MMMH | 15 | IP6K1 | 3 | 5.901e-08 | rs373797031 |
| ADHD | MMMH | 16 | CDHR4 | 3 | 4.367e-07 | rs373797031 |
| ADHD | MMMH | 17 | FAM212A | 3 | 2.557e-07 | rs373797031 |
| ADHD | MMMH | 18 | UBA7 | 3 | 4.367e-07 | rs373797031 |
| ADHD | MMMH | 19 | TRAIP | 3 | 5.619e-07 | rs373797031 |
| ADHD | MMMH | 20 | CAMKV | 3 | 9.356e-07 | rs373797031 |
| ADHD | MMMH | 21 | RBM6 | 3 | 1.998e-07 | rs373797031 |
| ADHD | MMMH | 22 | HYAL3 | 3 | 1.069e-07 | rs373797031 |
| ADHD | MMMH | 23 | MEF2C | 5 | 1.695e-07 | rs304136 |
| ADHD | MMMH | 24 | SPATS2 | 12 | 8.567e-06 | rs933738 |
| ADHD | MMMH | 25 | KCNH3 | 12 | 4.095e-07 | rs933738 |
| ADHD | MMMH | 26 | MCRS1 | 12 | 4.095e-07 | rs933738 |
| ADHD | MMMH | 27 | PRPF40B | 12 | 1.716e-06 | rs933738 |
| ADHD | MMMH | 28 | FAM186B | 12 | 1.716e-06 | rs933738 |
| ADHD | MMMH | 29 | FMNL3 | 12 | 7.508e-06 | rs933738 |
| ADHD | MMMH | 30 | TMBIM6 | 12 | 7.091e-06 | rs933738 |
| ADHD | MMMH | 31 | SH2B3 | 12 | 8.013e-07 | rs10849943 |
| ADHD | MMMH | 32 | ATXN2 | 12 | 3.011e-06 | rs10849943 |
| ADHD | MMMH | 33 | ALDH2 | 12 | 4.842e-07 | rs10849943 |
| ADHD | MMMH | 34 | TMEM116 | 12 | 9.193e-07 | rs10849943 |
| ADHD | MMMH | 35 | NAA25 | 12 | 4.842e-07 | rs10849943 |
| ADHD | MMMH | 36 | SERPINA1 | 14 | 4.065e-07 | rs112635299 |
| ADHD | MMMH | 37 | SEMA6D | 15 | 3.759e-07 | rs281287 |

**Suppl. Table 5.**
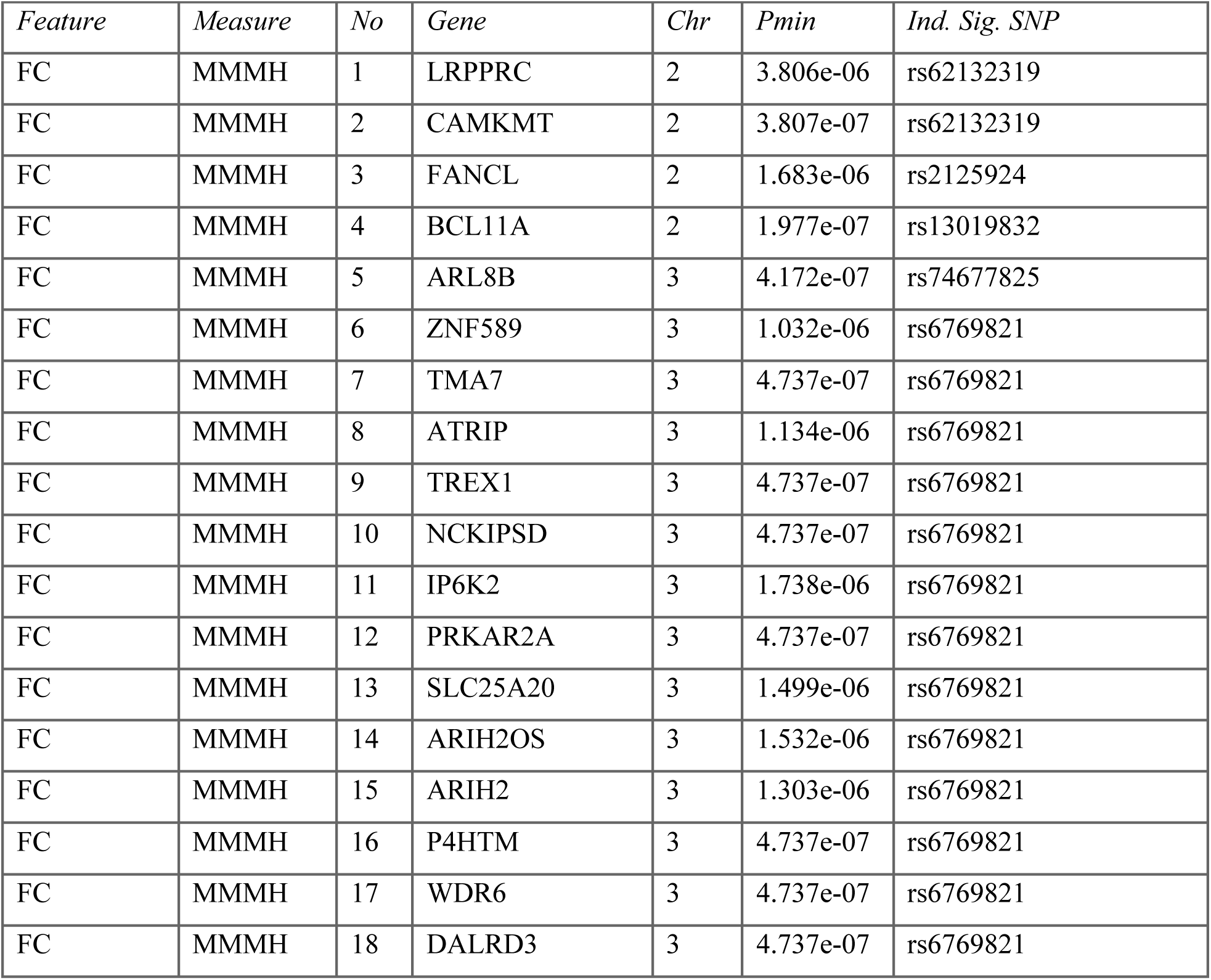
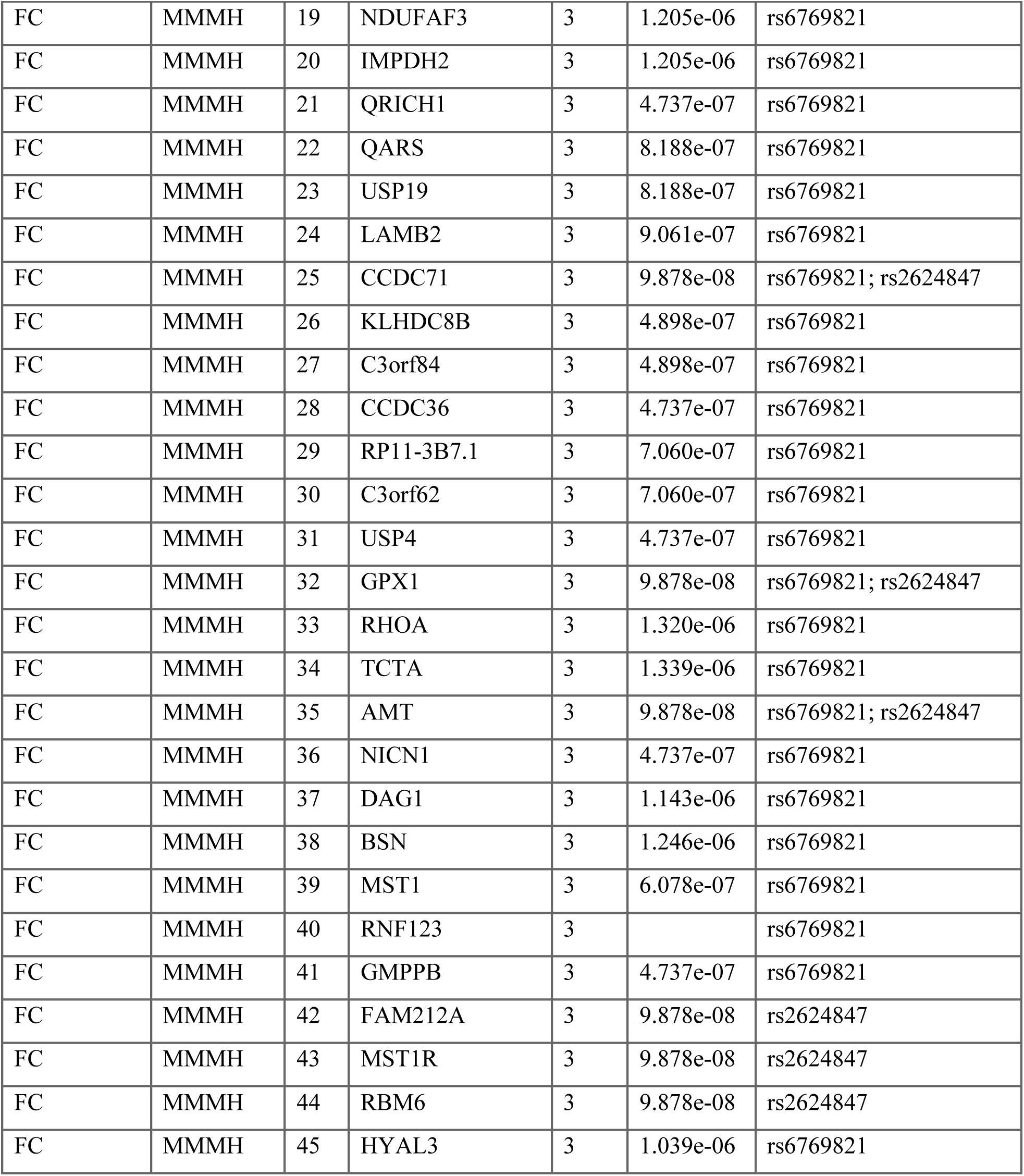
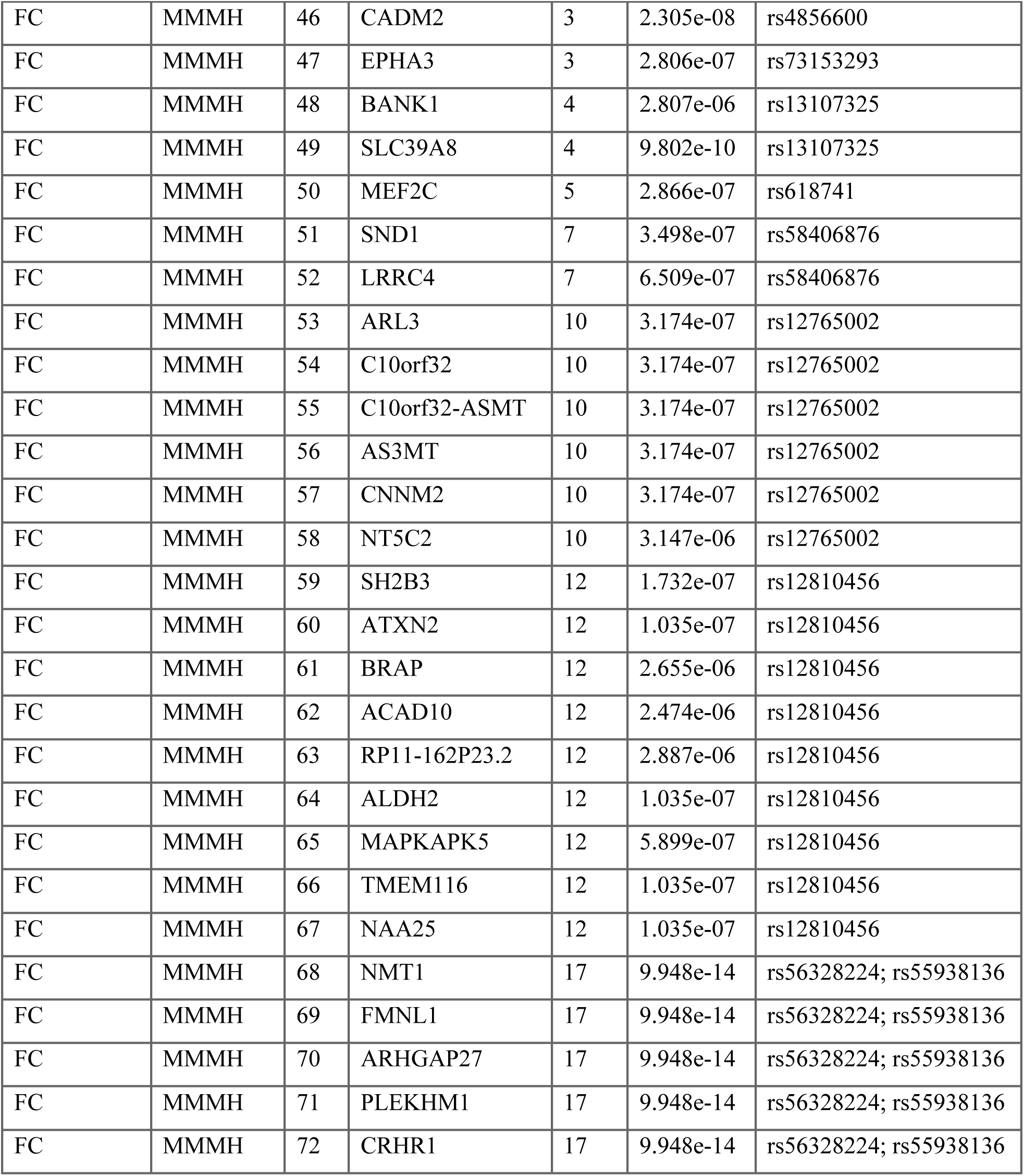
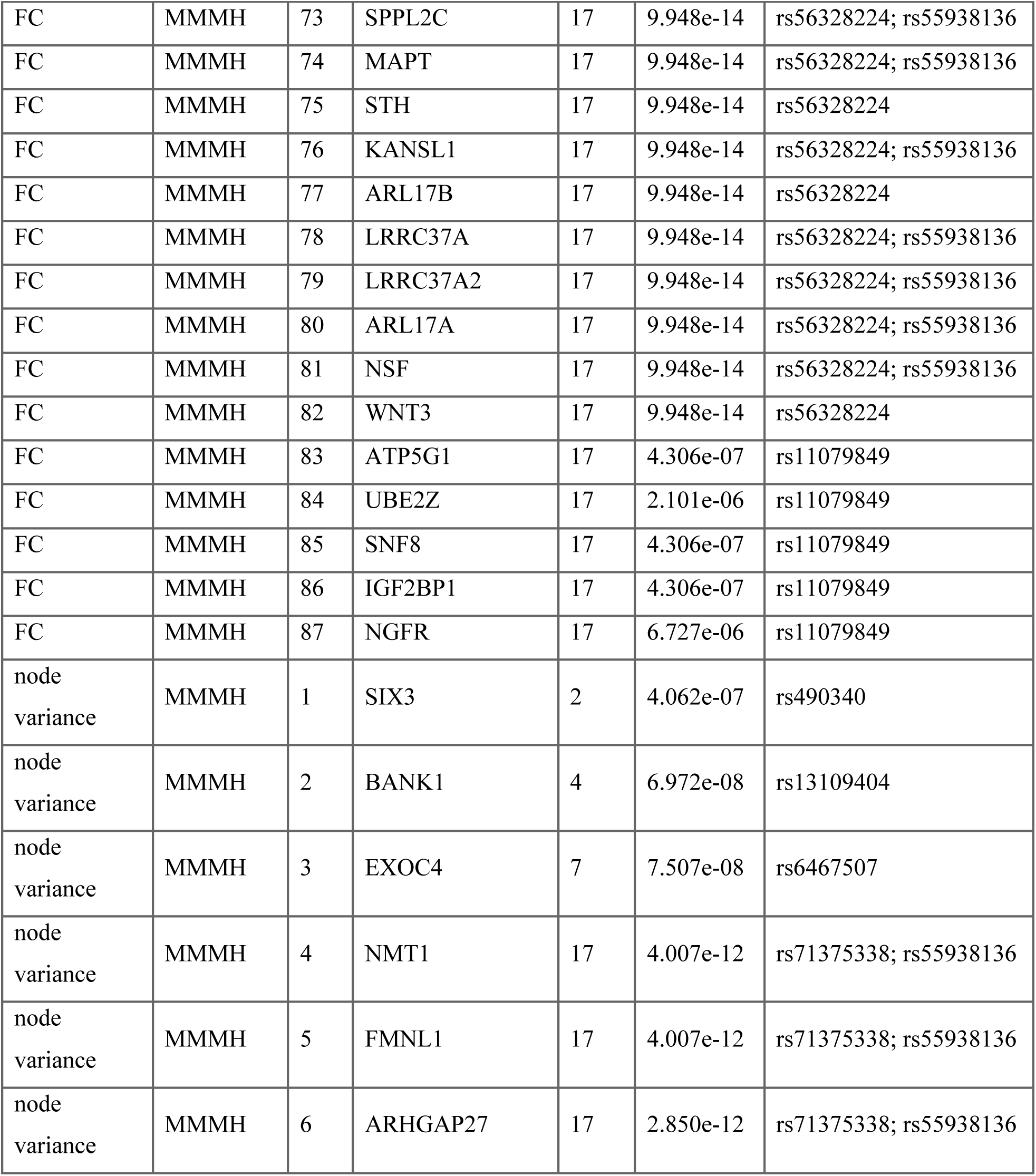
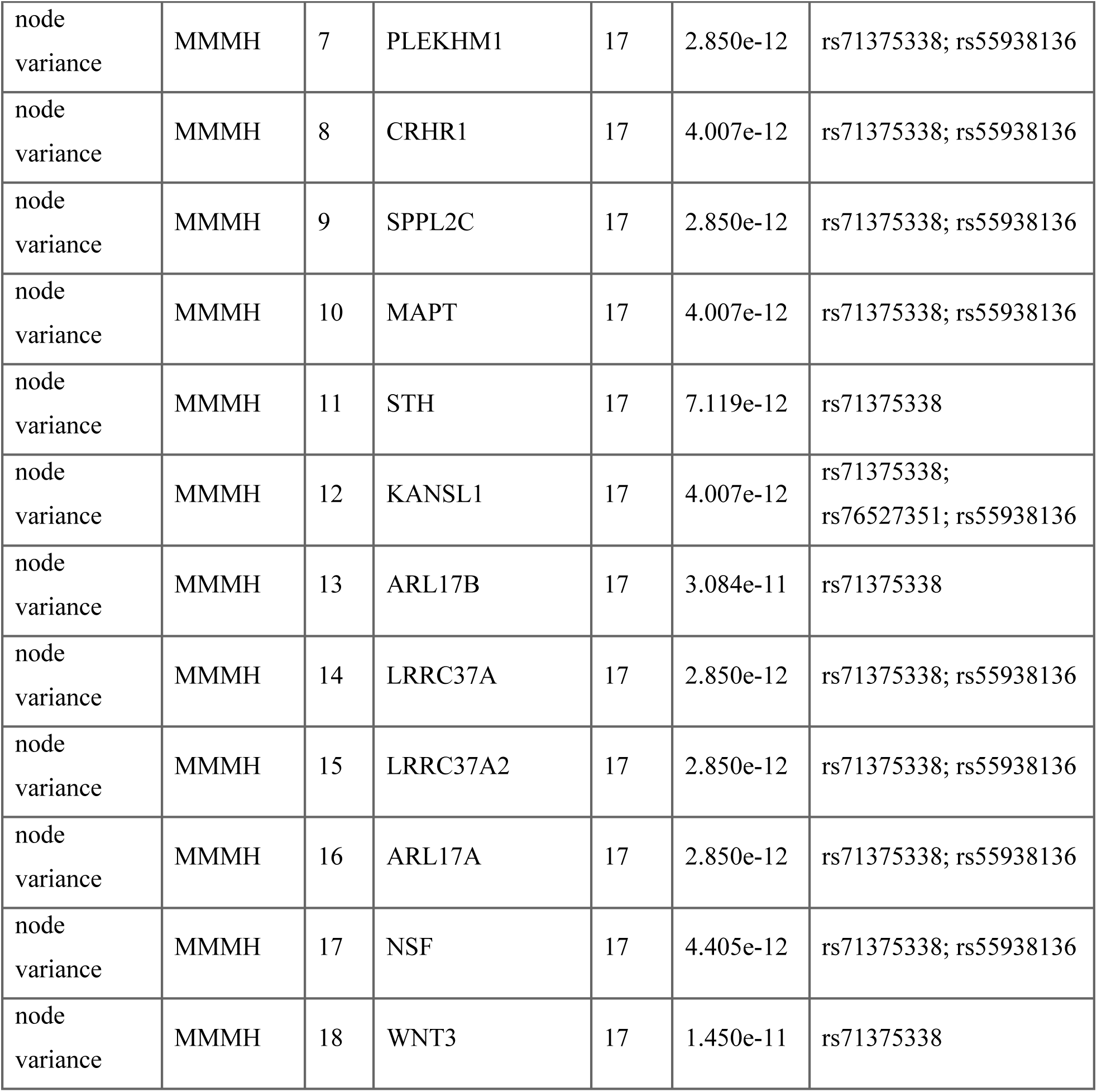
Gene list of conjFDR between the brain functional connectome and multivariate genome-wide association statistics for mental health. Genes identified through FUMA associated with the shared loci between the multivariate genome-wide association statistics for mental health and FC or node variance. The number of genes in this gene set can be larger than the number of loci since a locus can be associated more than one gene.

| *Feature* | *Measure* | *No* | *Gene* | *Chr* | *Pmin* | *Ind. Sig. SNP* |
| --- | --- | --- | --- | --- | --- | --- |
| FC | MMMH | 1 | LRPPRC | 2 | 3.806e-06 | rs62132319 |
| FC | MMMH | 2 | CAMKMT | 2 | 3.807e-07 | rs62132319 |
| FC | MMMH | 3 | FANCL | 2 | 1.683e-06 | rs2125924 |
| FC | MMMH | 4 | BCL11A | 2 | 1.977e-07 | rs13019832 |
| FC | MMMH | 5 | ARL8B | 3 | 4.172e-07 | rs74677825 |
| FC | MMMH | 6 | ZNF589 | 3 | 1.032e-06 | rs6769821 |
| FC | MMMH | 7 | TMA7 | 3 | 4.737e-07 | rs6769821 |
| FC | MMMH | 8 | ATRIP | 3 | 1.134e-06 | rs6769821 |
| FC | MMMH | 9 | TREX1 | 3 | 4.737e-07 | rs6769821 |
| FC | MMMH | 10 | NCKIPSD | 3 | 4.737e-07 | rs6769821 |
| FC | MMMH | 11 | IP6K2 | 3 | 1.738e-06 | rs6769821 |
| FC | MMMH | 12 | PRKAR2A | 3 | 4.737e-07 | rs6769821 |
| FC | MMMH | 13 | SLC25A20 | 3 | 1.499e-06 | rs6769821 |
| FC | MMMH | 14 | ARIH2OS | 3 | 1.532e-06 | rs6769821 |
| FC | MMMH | 15 | ARIH2 | 3 | 1.303e-06 | rs6769821 |
| FC | MMMH | 16 | P4HTM | 3 | 4.737e-07 | rs6769821 |
| FC | MMMH | 17 | WDR6 | 3 | 4.737e-07 | rs6769821 |
| FC | MMMH | 18 | DALRD3 | 3 | 4.737e-07 | rs6769821 |

|  |  |  |  |  |  |  |
| --- | --- | --- | --- | --- | --- | --- |
| *Feature* | *Measure* | *No* | *Gene* | *Chr* | *Pmin* | *Ind. Sig. SNP* |
| FC | MMMH | 19 | NDUFAF3 | 3 | 1.205e-06 | rs6769821 |
| FC | MMMH | 20 | IMPDH2 | 3 | 1.205e-06 | rs6769821 |
| FC | MMMH | 21 | QRICH1 | 3 | 4.737e-07 | rs6769821 |
| FC | MMMH | 22 | QARS | 3 | 8.188e-07 | rs6769821 |
| FC | MMMH | 23 | USP19 | 3 | 8.188e-07 | rs6769821 |
| FC | MMMH | 24 | LAMB2 | 3 | 9.061e-07 | rs6769821 |
| FC | MMMH | 25 | CCDC71 | 3 | 9.878e-08 | rs6769821; rs2624847 |
| FC | MMMH | 26 | KLHDC8B | 3 | 4.898e-07 | rs6769821 |
| FC | MMMH | 27 | C3orf84 | 3 | 4.898e-07 | rs6769821 |
| FC | MMMH | 28 | CCDC36 | 3 | 4.737e-07 | rs6769821 |
| FC | MMMH | 29 | RP11-3B7.1 | 3 | 7.060e-07 | rs6769821 |
| FC | MMMH | 30 | C3orf62 | 3 | 7.060e-07 | rs6769821 |
| FC | MMMH | 31 | USP4 | 3 | 4.737e-07 | rs6769821 |
| FC | MMMH | 32 | GPX1 | 3 | 9.878e-08 | rs6769821; rs2624847 |
| FC | MMMH | 33 | RHOA | 3 | 1.320e-06 | rs6769821 |
| FC | MMMH | 34 | TCTA | 3 | 1.339e-06 | rs6769821 |
| FC | MMMH | 35 | AMT | 3 | 9.878e-08 | rs6769821; rs2624847 |
| FC | MMMH | 36 | NICN1 | 3 | 4.737e-07 | rs6769821 |
| FC | MMMH | 37 | DAG1 | 3 | 1.143e-06 | rs6769821 |
| FC | MMMH | 38 | BSN | 3 | 1.246e-06 | rs6769821 |
| FC | MMMH | 39 | MST1 | 3 | 6.078e-07 | rs6769821 |
| FC | MMMH | 40 | RNF123 | 3 |  | rs6769821 |
| FC | MMMH | 41 | GMPPB | 3 | 4.737e-07 | rs6769821 |
| FC | MMMH | 42 | FAM212A | 3 | 9.878e-08 | rs2624847 |
| FC | MMMH | 43 | MST1R | 3 | 9.878e-08 | rs2624847 |
| FC | MMMH | 44 | RBM6 | 3 | 9.878e-08 | rs2624847 |
| FC | MMMH | 45 | HYAL3 | 3 | 1.039e-06 | rs6769821 |
| *Feature* | *Measure* | *No* | *Gene* | *Chr* | *Pmin* | *Ind. Sig. SNP* |
| FC | MMMH | 46 | CADM2 | 3 | 2.305e-08 | rs4856600 |
| FC | MMMH | 47 | EPHA3 | 3 | 2.806e-07 | rs73153293 |
| FC | MMMH | 48 | BANK1 | 4 | 2.807e-06 | rs13107325 |
| FC | MMMH | 49 | SLC39A8 | 4 | 9.802e-10 | rs13107325 |
| FC | MMMH | 50 | MEF2C | 5 | 2.866e-07 | rs618741 |
| FC | MMMH | 51 | SND1 | 7 | 3.498e-07 | rs58406876 |
| FC | MMMH | 52 | LRRC4 | 7 | 6.509e-07 | rs58406876 |
| FC | MMMH | 53 | ARL3 | 10 | 3.174e-07 | rs12765002 |
| FC | MMMH | 54 | C10orf32 | 10 | 3.174e-07 | rs12765002 |
| FC | MMMH | 55 | C10orf32-ASMT | 10 | 3.174e-07 | rs12765002 |
| FC | MMMH | 56 | AS3MT | 10 | 3.174e-07 | rs12765002 |
| FC | MMMH | 57 | CNNM2 | 10 | 3.174e-07 | rs12765002 |
| FC | MMMH | 58 | NT5C2 | 10 | 3.147e-06 | rs12765002 |
| FC | MMMH | 59 | SH2B3 | 12 | 1.732e-07 | rs12810456 |
| FC | MMMH | 60 | ATXN2 | 12 | 1.035e-07 | rs12810456 |
| FC | MMMH | 61 | BRAP | 12 | 2.655e-06 | rs12810456 |
| FC | MMMH | 62 | ACAD10 | 12 | 2.474e-06 | rs12810456 |
| FC | MMMH | 63 | RP11-162P23.2 | 12 | 2.887e-06 | rs12810456 |
| FC | MMMH | 64 | ALDH2 | 12 | 1.035e-07 | rs12810456 |
| FC | MMMH | 65 | MAPKAPK5 | 12 | 5.899e-07 | rs12810456 |
| FC | MMMH | 66 | TMEM116 | 12 | 1.035e-07 | rs12810456 |
| FC | MMMH | 67 | NAA25 | 12 | 1.035e-07 | rs12810456 |
| FC | MMMH | 68 | NMT1 | 17 | 9.948e-14 | rs56328224; rs55938136 |
| FC | MMMH | 69 | FMNL1 | 17 | 9.948e-14 | rs56328224; rs55938136 |
| FC | MMMH | 70 | ARHGAP27 | 17 | 9.948e-14 | rs56328224; rs55938136 |
| FC | MMMH | 71 | PLEKHM1 | 17 | 9.948e-14 | rs56328224; rs55938136 |
| FC | MMMH | 72 | CRHR1 | 17 | 9.948e-14 | rs56328224; rs55938136 |
| *Feature* | *Measure* | *No* | *Gene* | *Chr* | *Pmin* | *Ind. Sig. SNP* |
| FC | MMMH | 73 | SPPL2C | 17 | 9.948e-14 | rs56328224; rs55938136 |
| FC | MMMH | 74 | MAPT | 17 | 9.948e-14 | rs56328224; rs55938136 |
| FC | MMMH | 75 | STH | 17 | 9.948e-14 | rs56328224 |
| FC | MMMH | 76 | KANSL1 | 17 | 9.948e-14 | rs56328224; rs55938136 |
| FC | MMMH | 77 | ARL17B | 17 | 9.948e-14 | rs56328224 |
| FC | MMMH | 78 | LRRC37A | 17 | 9.948e-14 | rs56328224; rs55938136 |
| FC | MMMH | 79 | LRRC37A2 | 17 | 9.948e-14 | rs56328224; rs55938136 |
| FC | MMMH | 80 | ARL17A | 17 | 9.948e-14 | rs56328224; rs55938136 |
| FC | MMMH | 81 | NSF | 17 | 9.948e-14 | rs56328224; rs55938136 |
| FC | MMMH | 82 | WNT3 | 17 | 9.948e-14 | rs56328224 |
| FC | MMMH | 83 | ATP5G1 | 17 | 4.306e-07 | rs11079849 |
| FC | MMMH | 84 | UBE2Z | 17 | 2.101e-06 | rs11079849 |
| FC | MMMH | 85 | SNF8 | 17 | 4.306e-07 | rs11079849 |
| FC | MMMH | 86 | IGF2BP1 | 17 | 4.306e-07 | rs11079849 |
| FC | MMMH | 87 | NGFR | 17 | 6.727e-06 | rs11079849 |
| node variance | MMMH | 1 | SIX3 | 2 | 4.062e-07 | rs490340 |
| node variance | MMMH | 2 | BANK1 | 4 | 6.972e-08 | rs13109404 |
| node variance | MMMH | 3 | EXOC4 | 7 | 7.507e-08 | rs6467507 |
| node variance | MMMH | 4 | NMT1 | 17 | 4.007e-12 | rs71375338; rs55938136 |
| node variance | MMMH | 5 | FMNL1 | 17 | 4.007e-12 | rs71375338; rs55938136 |
| node variance | MMMH | 6 | ARHGAP27 | 17 | 2.850e-12 | rs71375338; rs55938136 |
| node variance | MMMH | 7 | PLEKHM1 | 17 | 2.850e-12 | rs71375338; rs55938136 |
| node variance | MMMH | 8 | CRHR1 | 17 | 4.007e-12 | rs71375338; rs55938136 |
| node variance | MMMH | 9 | SPPL2C | 17 | 2.850e-12 | rs71375338; rs55938136 |
| node variance | MMMH | 10 | MAPT | 17 | 4.007e-12 | rs71375338; rs55938136 |
| node variance | MMMH | 11 | STH | 17 | 7.119e-12 | rs71375338 |
| node variance | MMMH | 12 | KANSL1 | 17 | 4.007e-12 | rs71375338;<br>rs76527351; rs55938136 |
| node variance | MMMH | 13 | ARL17B | 17 | 3.084e-11 | rs71375338 |
| node variance | MMMH | 14 | LRRC37A | 17 | 2.850e-12 | rs71375338; rs55938136 |
| node variance | MMMH | 15 | LRRC37A2 | 17 | 2.850e-12 | rs71375338; rs55938136 |
| node variance | MMMH | 16 | ARL17A | 17 | 2.850e-12 | rs71375338; rs55938136 |
| node variance | MMMH | 17 | NSF | 17 | 4.405e-12 | rs71375338; rs55938136 |
| node variance | MMMH | 18 | WNT3 | 17 | 1.450e-11 | rs71375338 |

**Suppl. Table 6.** Biological processes mapped by reactome. Biological processes associated with the shared genetic determinants between the multivariate genome-wide association statistics for mental health and the brain functional connectome as identified by the reactome toolbox.

|  |  |  |  |  |
| --- | --- | --- | --- | --- |
| *Pathway name* | *No of genes involved* | *p-value* | *p-value (FDR)* | *Genes involved* |
| Axonal growth stimulation | 2 | 0.0005 | 0.2235 | NGFR; RHOA |
| Signaling by MST1 | 2 | 0.0014 | 0.2491 | MST1; MST1R |
| MECP2 regulates transcription factors | 2 | 0.0021 | 0.2491 | MEF2C |
| Axonal growth inhibition (RHOA activation) | 2 | 0.0026 | 0.2491 | NGFR; RHOA |
| p75NTR regulates axonogenesis | 2 | 0.0030 | 0.2491 | NGFR; RHOA |
| Respiratory electron transport, ATP synthesis by chemiosmotic coupling, and heat production by uncoupling proteins. | 5 | 0.0038 | 0.2595 | NDUFAF3;<br>QRICH1;<br>ATP5G1;<br>LRPPRC |
| Inactivation, recovery and regulation of the phototransduction cascade | 3 | 0.0059 | 0.3361 | NMT1;<br>CAMKMT |
| The phototransduction cascade | 3 | 0.0075 | 0.3361 | NMT1;<br>CAMKMT |
| Non-integrin membrane-ECM interactions | 3 | 0.0082 | 0.3361 | LAMB2; DAG1 |
| Formation of ATP by chemiosmotic coupling | 2 | 0.0106 | 0.3361 | ATP5G1 |
| ECM proteoglycans | 3 | 0.0164 | 0.3361 | LAMB2; DAG1 |
| Laminin interactions | 2 | 0.0187 | 0.3361 | LAMB2 |
| Cristae formation | 2 | 0.0187 | 0.3361 | ATP5G1 |
| MET activates PTK2 signaling | 2 | 0.0198 | 0.3361 | LAMB2 |
| NFG and proNGF binds to p75NTR | 1 | 0.0199 | 0.3361 | NGFR |
| Defective POMGNT1 causes MDDGA3, MDDGB3 and MDDGC3 | 1 | 0.0199 | 0.3361 | DAG1 |
| EPHA-mediated growth cone collapse | 2 | 0.0210 | 0.3361 | EPHA3; RHOA |
| The citric acid (TCA) cycle and respiratory electron transport | 5 | 0.0224 | 0.3361 | NDUFAF3;<br>QRICH1;<br>ATP5G1;<br>LRPPRC |
| *Pathway name* | *No of genes involved* | *p-value* | *p-value (FDR)* | *Genes involved* |
| Defective POMT2 causes MDDGA2, MDDGB2 and MDDGC2 | 1 | 0.0265 | 0.3361 | DAG1 |
| Defective POMT1 causes MDDGA1, MDDGB1 and MDDGC1 | 1 | 0.0265 | 0.3361 | DAG1 |
| Activation, myristoylation of BID and translocation to mitochondria | 1 | 0.0265 | 0.3361 | NMT1 |
| EGR2 and SOX10-mediated initiation of Schwann cell myelination | 2 | 0.0286 | 0.3361 | LAMB2; DAG1 |
| MAPK6/MAPK4 signaling | 3 | 0.0349 | 0.3361 | IGF2BP1; MAPKAPK5 |
| MET promotes cell motility | 2 | 0.0371 | 0.3361 | LAMB2 |
| NADE modulates death signalling | 1 | 0.0394 | 0.3361 | NGFR |
| p75NTR negatively regulates cell cycle via SC1 | 1 | 0.0394 | 0.3361 | NGFR |
| Cytosolic tRNA aminoacylation | 2 | 0.0448 | 0.3361 | QARS |
| Respiratory electron transport | 3 | 0.0454 | 0.3361 | NDUFAF3; QRICH1; LRPPRC |
| Regulation by TREX1 | 1 | 0.0458 | 0.3361 | TREX1 |
| Trafficking of myristoylated proteins to the cilium | 1 | 0.0458 | 0.3361 | ARL3 |
| Fanconi Anemia Pathway | 2 | 0.0497 | 0.3361 | FANCL; ATRIP |

## Notes

### Author Declarations

All data used in this study are part of the publicly available UK Biobank initiative (https://www.ukbiobank.ac.uk/).

## References

1. Pardiñas, A. F. et al. Common schizophrenia alleles are enriched in mutation-intolerant genes and in regions under strong background selection. Nat. Genet. 50, 381–389 (2018).

2. Pettersson-Yeo, W., Allen, P., Benetti, S., McGuire, P. & Mechelli, A. Dysconnectivity in schizophrenia: where are we now? Neurosci. Biobehav. Rev. 35, 1110–1124 (2011).

3. Trubetskoy, V. et al. Mapping genomic loci implicates genes and synaptic biology in schizophrenia. Nature 604, 502–508 (2022).

4. Mullins, N. et al. Genome-wide association study of more than 40,000 bipolar disorder cases provides new insights into the underlying biology. Nat. Genet. 53, 817–829 (2021).

5. Vai, B., Bertocchi, C. & Benedetti, F. Cortico-limbic connectivity as a possible biomarker for bipolar disorder: where are we now? Expert Rev. Neurother. 19, 159–172 (2019).

6. Wray, N. R. et al. Genome-wide association analyses identify 44 risk variants and refine the genetic architecture of major depression. Nat. Genet. 50, 668–681 (2018).

7. Zhuo, C. et al. The rise and fall of MRI studies in major depressive disorder. Transl. Psychiatry 9, 335 (2019).

8. Otowa, T. et al. Meta-analysis of genome-wide association studies of anxiety disorders. Mol. Psychiatry 21, 1391–1399 (2016).

9. Stein, M. B. Neurobiology of generalized anxiety disorder. J. Clin. Psychiatry 70 Suppl 2, 15–19 (2009).

10. Thompson, P. M. et al. ENIGMA and global neuroscience: A decade of large-scale studies of the brain in health and disease across more than 40 countries. Transl. Psychiatry 10, 100 (2020).

11. Cheng, W. et al. Genetic Association Between Schizophrenia and Cortical Brain Surface Area and Thickness. JAMA Psychiatry 78, 1020–1030 (2021).

12. Smeland, O. B. et al. Genetic Overlap Between Schizophrenia and Volumes of Hippocampus, Putamen, and Intracranial Volume Indicates Shared Molecular Genetic Mechanisms. Schizophr. Bull. 44, 854–864 (2018).

13. Roelfs, D. et al. Genetic overlap between multivariate measures of human functional brain connectivity and psychiatric disorders. medRxiv 2021.06.15.21258954 (2021) doi:10.1101/2021.06.15.21258954.

14. Anttila, V. et al. Analysis of shared heritability in common disorders of the brain. Science 360, eaap8757 (2018).

15. Bulik-Sullivan, B. et al. An atlas of genetic correlations across human diseases and traits. Nat Genet 47, 1236–41 (2015).

16. Cross-Disorder Group of the Psychiatric Genomics Consortium. Identification of risk loci with shared effects on five major psychiatric disorders: a genome-wide analysis. The Lancet 381, 1371–1379 (2013).

17. Lee, P. H. et al. Genomic Relationships, Novel Loci, and Pleiotropic Mechanisms across Eight Psychiatric Disorders. Cell 179, 1469-1482.e11 (2019).

18. Romero, C. et al. Exploring the genetic overlap between 12 psychiatric disorders. MedRxiv 2022.04.12.22273763 (2022) doi:10.1101/2022.04.12.22273763.

19. Wardenaar, K. J. & de Jonge, P. Diagnostic heterogeneity in psychiatry: towards an empirical solution. BMC Med. 11, 201 (2013).

20. Widiger, T. A. & Clark, L. A. Toward DSM—V and the classification of psychopathology. Psychol. Bull. 126, 946–963 (2000).

21. Kessler, R. C. et al. Lifetime Prevalence and Age-of-Onset Distributions of DSM-IV Disorders in the National Comorbidity Survey Replication. Arch. Gen. Psychiatry 62, 593–602 (2005).

22. McGrath, J. J. et al. Psychotic Experiences in the General Population: A Cross-National Analysis Based on 31,261 Respondents From 18 Countries. JAMA Psychiatry 72, 697–705 (2015).

23. Doherty, J. L. & Owen, M. J. Genomic insights into the overlap between psychiatric disorders: implications for research and clinical practice. Genome Med. 6, 29–29 (2014).

24. Roelfs, D. et al. Phenotypically independent profiles relevant to mental health are genetically correlated. Transl. Psychiatry 11, 202 (2021).

25. van der Meer, D. et al. Understanding the genetic determinants of the brain with MOSTest. Nat. Commun. 11, 3512 (2020).

26. Sudlow, C. et al. UK biobank: an open access resource for identifying the causes of a wide range of complex diseases of middle and old age. PLoS Med 12, e1001779 (2015).

27. Alfaro-Almagro, F. et al. Image processing and Quality Control for the first 10,000 brain imaging datasets from UK Biobank. NeuroImage 166, 400–424 (2018).

28. Watanabe, K., Taskesen, E., van Bochoven, A. & Posthuma, D. Functional mapping and annotation of genetic associations with FUMA. Nat. Commun. 8, 1826 (2017).

29. de Leeuw, C. A., Mooij, J. M., Heskes, T. & Posthuma, D. MAGMA: Generalized Gene-Set Analysis of GWAS Data. PLOS Comput. Biol. 11, e1004219 (2015).

30. Jassal, B. et al. The reactome pathway knowledgebase. Nucleic Acids Res. 48, D498–D503 (2020).

31. Andreassen, O. A. et al. Improved detection of common variants associated with schizophrenia and bipolar disorder using pleiotropy-informed conditional false discovery rate. PLoS Genet 9, e1003455 (2013).

32. Demontis, D. et al. Discovery of the first genome-wide significant risk loci for attention deficit/hyperactivity disorder. Nat. Genet. 51, 63–75 (2019).

33. Grove, J. et al. Identification of common genetic risk variants for autism spectrum disorder. Nat. Genet. 51, 431–444 (2019).

34. Duncan, L. E. et al. Largest GWAS of PTSD (N=20 070) yields genetic overlap with schizophrenia and sex differences in heritability. Mol. Psychiatry 23, 666–673 (2018).

35. Smeland, O. B. et al. Discovery of shared genomic loci using the conditional false discovery rate approach. Hum. Genet. 139, 85–94 (2020).

36. Visscher, P. M. et al. 10 Years of GWAS Discovery: Biology, Function, and Translation. Am. J. Hum. Genet. 101, 5–22 (2017).

37. Devor, A. et al. Genetic evidence for role of integration of fast and slow neurotransmission in schizophrenia. Mol. Psychiatry 22, 792–801 (2017).

38. Hsu, W.-C. J., Nilsson, C. L. & Laezza, F. Role of the axonal initial segment in psychiatric disorders: function, dysfunction, and intervention. Front. Psychiatry 5, 109–109 (2014).

39. Mukai, J. et al. Molecular substrates of altered axonal growth and brain connectivity in a mouse model of schizophrenia. Neuron 86, 680–695 (2015).

40. Dutt, R. K. et al. Mental health in the UK Biobank: A roadmap to self-report measures and neuroimaging correlates. Hum. Brain Mapp. 43, 816–832 (2022).

41. Lyall, D. M. et al. Quantifying bias in psychological and physical health in the UK Biobank imaging sub-sample. Brain Commun. 4, fcac119 (2022).

42. Stahl, E. A. et al. Genome-wide association study identifies 30 loci associated with bipolar disorder. Nat. Genet. 51, 793–803 (2019).

43. Duncan, L. E. et al. Largest GWAS of PTSD (N=20 070) yields genetic overlap with schizophrenia and sex differences in heritability. Mol. Psychiatry 23, 666–673 (2018).

